# Effects of High-Intensity Interval Training on Cardiorespiratory Fitness in Patients with Stroke: A Meta-Analysis

**DOI:** 10.1101/2025.09.29.25336695

**Authors:** Seo Hyun Kim, Min-Su Kim

## Abstract

**BACKGROUND:** Stroke survivors typically exhibit impaired cardiorespiratory fitness (CRF) and walking ability, which hinder their recovery and independence. High-intensity interval training (HIIT) has been proposed as a time-efficient strategy, but its efficacy and safety remain limited.

**METHODS:** We conducted a systematic review and meta-analysis in accordance with the Preferred Reporting Items for Systematic Reviews and Meta-Analysis (PRISMA) guidelines. MEDLINE, EMBASE, Web of Science, and Cochrane CENTRAL were searched (January 2025–July 2025) for randomized controlled trials (RCTs) comparing HIIT with low- to moderate-intensity training or conventional rehabilitation in stroke survivors. The primary outcomes were CRF and walking ability, and subgroup analyses were conducted for patient characteristics and intervention dose.

**RESULTS:** Eleven randomized controlled trials (551 participants) of moderate-to-high quality were included. Compared with conventional rehabilitation, HIIT significantly improved CRF (MD 3.12 mL/kg/min; 95% CI 2.15–4.09; I^2^ = 85%) and walking ability (MD 0.11 m/s; 95% CI 0.03–0.19; I^2^ = 77%); these values exceeded clinically meaningful thresholds. Subgroup analyses indicated greater improvements in CRF among middle-aged participants (≤ 60 years; MD 4.18 mL/kg/min), and the benefits were consistent across stroke severity and chronicity. Although intervention times exceeding 20 minutes provided additional gains, no significant differences were observed in terms of intervention frequency or program length.

**CONCLUSIONS:** Compared with conventional rehabilitation, HIIT improves cardiorespiratory fitness and walking ability in stroke survivors, thus identifying this approach as an effective and time-efficient strategy.

**REGISTRATION:** URL: https://www.crd.york.ac.uk/PROSPERO/; Unique identifier: CRD420251111351.

## Introduction

Stroke is a leading cause of disability worldwide and is the third most common cause of death.^1^ In 2019, approximately 6.5 million people died from stroke, and the global burden was estimated at 143 million disability-adjusted life years (DALYs) lost.^2^ The increasing burden of stroke-related mortality and disability can largely be attributed to the high prevalence of modifiable cardiovascular risk factors, reduced cardiorespiratory fitness (CRF), and impaired walking ability among stroke survivors.^3–5^ These factors contribute to physical inactivity and deconditioning, which represent major barriers to full participation in rehabilitation programs and attenuate potential motor recovery.^6,7^ Moreover, this situation leads to a vicious cycle of deconditioning, an increased risk of recurrent stroke, and greater cardiovascular morbidity.^8^

CRF reflects the ability of the circulatory and respiratory systems to deliver oxygen to skeletal muscle during sustained, moderate-to high-intensity physical activity. After stroke, CRF is impaired, with peak oxygen uptake (VO_2_peak) values ranging from 8–22 mL/kg/min, which is equivalent to 26% to 87% of those observed in age- and sex-matched healthy individuals.^9^ Walking ability is also substantially compromised. Previous studies have reported that 22% of stroke survivors are unable to walk, 14% require walking aids, and fewer than 10% achieve a walking speed sufficient to regain independence and participate in community _life.10,11_

To address these limitations, aerobic exercise is strongly recommended for improving CRF and managing cardiovascular risk factors, with moderate-intensity continuous training (MICT) being the most commonly prescribed modality.^12^ Since the American Heart Association guidelines were released in 2007, however, there has been growing clinical interest in higher-intensity aerobic training.^13^ Recent systematic reviews have shown that high-intensity aerobic exercise result in greater improvements in CRF and physical function among individuals with stroke.^14,15^ High-intensity continuous training has shown efficacy in enhancing CRF and walking performance, but its applicability is restricted in patients with low baseline fitness, motor impairments, or weakness.^16,17^

Consequently, high-intensity interval training (HIIT) has emerged as a time-efficient and effective alternative to traditional continuous aerobic exercise. HIIT consists of repeated bouts of near-maximal intensity exercise interspersed with periods of low-intensity activity or rest. A growing body of systematic reviews and meta-analyses has supported the beneficial effects and safety of HIIT in both healthy adults and individuals with chronic diseases.^14,15,18,19^ Recent systematic reviews have reported that high-intensity exercise improve CRF among stroke survivors and may represent a safe intervention for poststroke rehabilitation.^14,15^ However, those reviews included nonrandomized studies in their meta-analyses, and in some cases, studies that did not implement HIIT as an intervention were analyzed. In addition, the comparators varied widely, ranging from home-based nonexercise education to MICT, making it difficult to clearly determine whether high-intensity training is more effective than established interventions are.

Therefore, in this study, a systematic review and meta-analysis restricted to randomized controlled trials (RCTs) were conducted to investigate the effects of HIIT on CRF and walking ability in patients after stroke, with additional analyses considering patient characteristics (age, stroke stage, and severity), intervention dose (session time, frequency, and duration), and safety outcomes. Furthermore, we assessed the safety of HIIT as a rehabilitation strategy for patients after stroke.

## Methods

This systematic review and meta-analysis was conducted and reported in accordance with the Preferred Reporting Items for Systematic Review and Meta-analysis (PRISMA) guidelines. The study protocol was prospectively registered in PROSPERO (registration number: CRD420251111351).

### Definitions

HIIT was defined as repeated short bouts of vigorous exercise performed at maximal or near-maximal effort interspersed with periods of low-intensity activity or rest. In this review, HIIT was considered exercise performed at ≥60% heart rate reserve (HRR) or VO_2_peak, ≥70% of maximal heart rate, or a Borg Ratings of Perceived Exertion (RPE) score ≥14.^14^ The classification of relative exercise intensity, adapted from the American College of Sports Medicine, is presented in Table 1.

**Table 1.** American College of Sports Medicine Classification of Exercise Relative Intensity.

| Intensity | %HRR or VO<sub>2</sub>R | %peak VO<sub>2</sub> | %peak HR | RPE Borg Scale |
| --- | --- | --- | --- | --- |
| Very light | <20 | <25 | <35 | <10 |
| Light | 20-39 | 25-44 | 35-54 | 10-11 |
| Moderate | 40-59 | 45-59 | 55-69 | 12-13 |
| Heavy | 60-84 | 60-84 | 70-89 | 14-16 |
| Very heavy | ≥85 | ≥85 | ≥90 | 17-19 |
| Maximal | 100 | 100 | 100 | 20 |

HRR indicates heart rate reserve; VO<sub>2</sub>R, VO<sub>2</sub> reserve; HR, heart rate; and RPE, rating of perceived exertion.

CRF was evaluated using peak oxygen uptake (VO_2_peak), or maximal oxygen uptake, which refers to oxygen consumption capacity during progressive exercise and reflects maximal cardiopulmonary capacity.^20^ VO_2_peak was generally measured using breath-by-breath analysis with a tightly fitted face mask, volume flow sensor, and gas analyzers for oxygen and carbon dioxide.^21^

Walking ability was assessed via gait speed, a reliable and valid indicator of lower limb function and general physical performance in stroke survivors.^22^ Higher gait speed indicates better physical function and walking ability. Gait speed was typically measured by timing participants as they walked over a set distance—usually 10 m.

For stroke stage, onset time was classified into subacute (≤6 months) and chronic (>6 months). Stroke severity was categorized into mild and moderate according to clinical assessments such as the National Institute of Health Stroke Scale, modified Barthel index, and Fugl–Meyer assessment.

### Search Strategy

A systematic search for studies was performed in the MEDLINE database via PubMed, EMBASE, Web of Science, and the Cochrane Central Register of Controlled Trials published from January 2005 to June 2025. Each source was searched on the same date, with the last search performed on July 17, 2025. Search terms combined keywords related to the population (e.g., “stroke” and “cerebrovascular accident”), intervention (e.g., “high-intensity training” and “aerobic interval exercise”), comparator (e.g., “moderate exercise”, “continuous physical activity”, and “rehabilitation program”), and outcomes (e.g., “cardiorespiratory fitness” and “gait speed”). The detailed search terms and combination methods are presented in the Supplemental Material.

After eliminating duplicates, two investigators (S.K. and M.K.) independently screened the titles and abstracts of the retrieved articles to accurately identify studies relevant to the research topic. Discrepancies were resolved through discussion and consensus. Furthermore, reference lists from the included studies were examined to identify additional relevant records. The full texts of potentially relevant articles were reviewed by two investigators (S.K. and M.K.), and any disagreements were resolved by collaborative consensus. This process was entirely manual, and no automated or artificial intelligence tools were used.

### Inclusion and Exclusion Criteria

Eligible studies were RCTs that provided a detailed description of the intervention, including exercise intensity, duration, and frequency, with the target intensity meeting at least one of the following criteria: ≥60% HRR, ≥70% peak HR, ≥14 on the Borg scale, or maximal walking/running speed. The control group was required to receive low-to moderate-intensity exercise (<60% HRR, <70% peak HR, <14 Borg scale), conventional physical therapy, or standard care. The Included studies reported at least one outcome used to assess CRF or walking ability.

Studies that reported the effects of high-intensity training without interval components were excluded. Studies were excluded if the control group did not receive a specific exercise intervention or if relevant outcome data were unavailable. Only full-text articles published in English were included, and conference abstracts, case reports, observational studies, and studies with insufficient data were excluded.

### Risk of Bias and Certainty of Evidence

Two reviewers (S.K. and M.K.) independently assessed methodological quality using the Physiotherapy Evidence Database (PEDro) scale (0–10). This validated tool, widely used in exercise and rehabilitation research, includes 11 items scored as “yes” (1) or “no” (0), with a maximum possible score of 10. RCTs were categorized as poor (<4), moderate (4–5), or high quality (6–10). Only studies with a PEDro score ≥4 (moderate to high quality) were included.^14^

In addition, the certainty of the evidence regarding the primary outcomes (VO_2_peak and gait speed) was evaluated using the Grading of Recommendations, Assessment, Development and Evaluation (GRADE) approach.^23^ This method rates evidence as high, moderate, low, or very low by considering five domains: risk of bias, inconsistency, indirectness, imprecision, and publication bias.

### Data Extraction and Synthesis

Data were extracted from all eligible studies using a Microsoft Excel spreadsheet developed for this review by the primary reviewer (S.K.) and cross-checked by the secondary reviewer (M.K.). Uncertainty was resolved by discussion and consensus. The data extracted included publication information (author details, journal information, year published, country, and design), participant characteristics (number of participants, mean age, time since stroke, and severity of stroke), intervention characteristics (intervention type and dose [frequency, intensity, and time]), comparison intervention characteristics, risks/adverse events, and results (VO_2_peak and gait speed).

When studies reported means and 95% confidence intervals (CIs) without standard deviations (SDs), SDs were estimated from the CI using the method described in the Cochrane Handbook. In cases where gait speed was not directly provided but distance from a 6-minute walk test was available, gait speed was calculated by dividing the distance (m) by 360 seconds. Other unit conversions were performed to ensure consistency across studies.

Adverse events were also extracted when reported by the included trials to provide an overview of the safety of HIIT. Because adverse events were inconsistently reported and often lacked sufficient detail for quantitative pooling, no meta-analysis was performed. Instead, all available adverse event data were narratively summarized, including the type of events as described by the original authors.

### Statistical Analysis

REX software (ver. 3.0.3; Rexsoft Inc., Seoul, Korea) was used to cunduct the meta-analysis. This meta-analysis included each intervention for which at least 2 included studies with comparable outcomes were identified. For all studies that reported continuous data, we chose the standardized mean difference (SMD) with a 95% CI as the summary statistic for the meta-analysis. The DerSimonian‒Laird method was used to calculate the random-effects meta-analytic estimate of the SMD for pre-post intervention variations in VO_2_peak and gait speed between the experimental and control groups. We used the I^2^ test to assess the statistical heterogeneity of the intervention effect among studies. I^2^ values of 25%, 50%, and 75% were considered low, moderate, and high heterogeneity, respectively. The findings are presented as forest plots.

Six analyses with subgroups were planned: participant characteristics (age [≤ 60 years vs. > 60 years], stroke stage [subacute vs. chronic], stroke severity [mild vs. moderate]) and intervention dose (time [≤20 minutes vs. >20 minutes], frequency [≤3 days a week vs. >3 days a week], and length [≤8 weeks vs. >8 weeks]) in the experimental group.

Publication bias was evaluated using a funnel plot and Egger’s test, with *P*<0.1 defined as indicating significant publication bias. To assess the robustness of the synthesized results and explore the potential impact of publication bias, we conducted a sensitivity analysis using the Copas selection model. This model evaluates how the estimated treatment effect changes under different assumptions about the probability of study selection on the basis of study size and effect size. An analysis of the primary outcomes (VO_2_peak and gait speed) was performed, and the results were compared with those obtained from the main random-effects meta-analysis.

## Results

### Search Results

The study selection process is summarized in Figure 1. A total of 11 studies, including 551 participants, were eligible for inclusion in the meta-analysis.^16,21,24–32^ The initial database search yielded 457 records, 124 of which were duplicates. An additional 5 studies were identified through reference list screening. After screening the titles and abstracts, 32 full texts were reviewed. Of these, 21 did not meet the inclusion criteria; the remaining 11 studies were included in the analysis. The characteristics of the included studies are summarized in Table 2.

**Figure 1.**
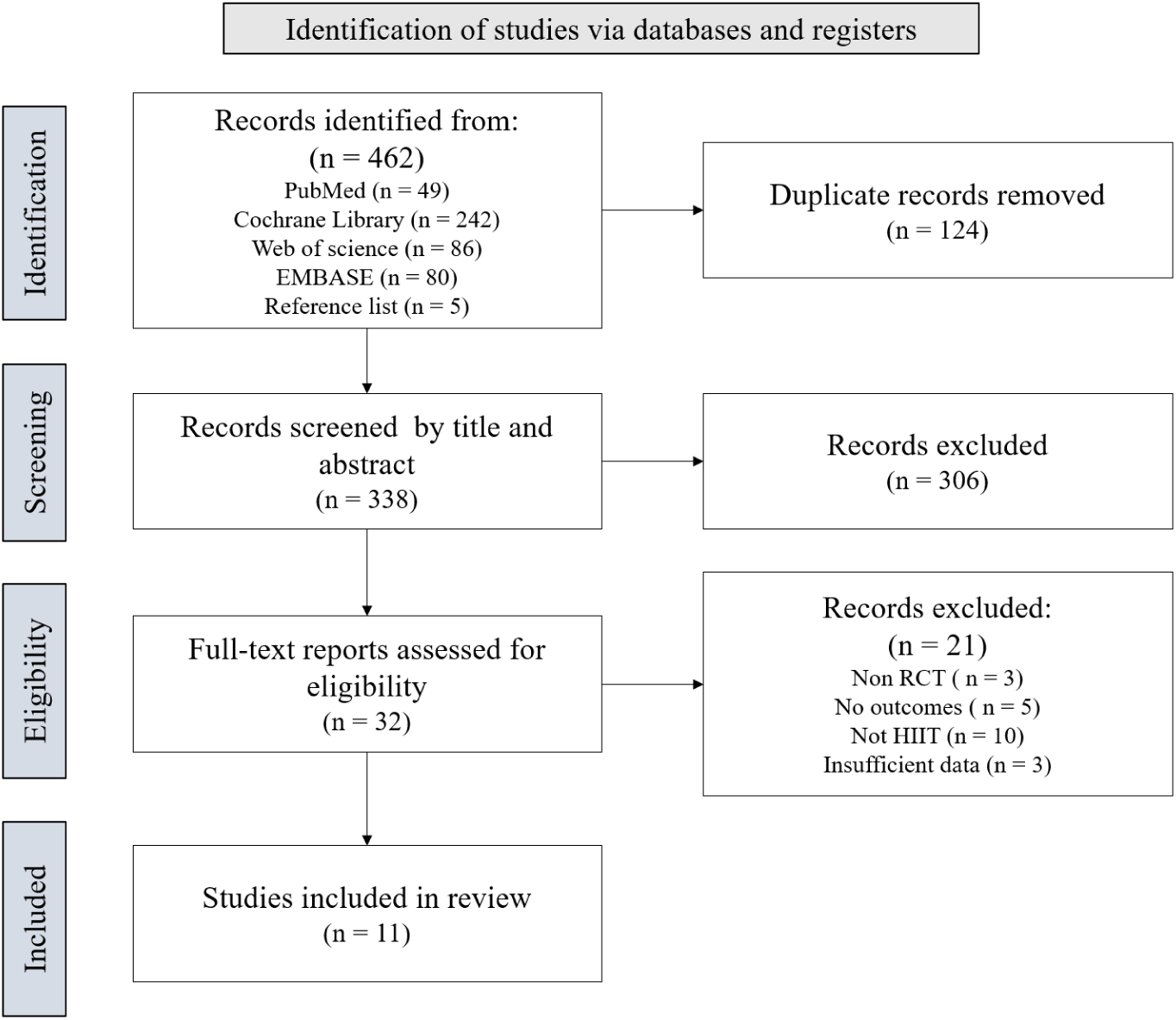
Flow Chart of the Search Process. The diagram illustrates the identification, screening, eligibility assessment, and inclusion of studies in accordance with the PRISMA guidelines.

**Table 2.**
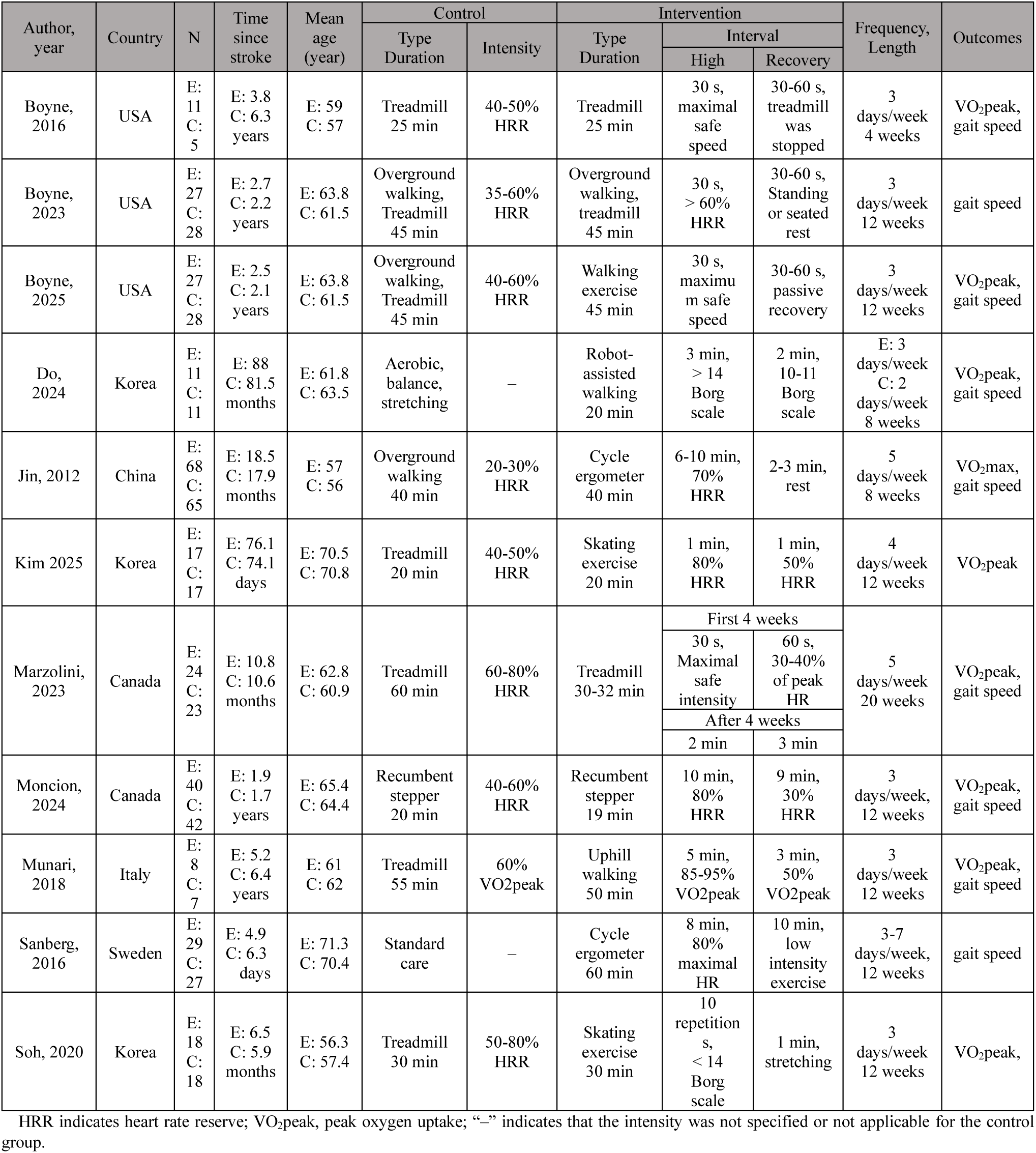
Summary of Studies Examining the Effects of HIIT on Patients with Stroke.

| Author,<br>year | Country | N | Time<br>since<br>stroke | Mean<br>age<br>(year) | Control |  | Intervention |  |  | Frequency,<br>Length | Outcomes |
| --- | --- | --- | --- | --- | --- | --- | --- | --- | --- | --- | --- |
|  |  |  |  |  | Type<br>Duration | Intensity | Type<br>Duration | Interval |  |  |  |
|  |  |  |  |  |  |  |  | High | Recovery |  |  |
| Boyne,<br>2016 | USA | E: 11<br>C: 5 | E: 3.8<br>C: 6.3<br>years | E: 59<br>C: 57 | Treadmill<br>25 min | 40-50%<br>HRR | Treadmill<br>25 min | 30 s,<br>maximal<br>safe<br>speed | 30-60 s,<br>treadmill<br>was<br>stopped | 3<br>days/week<br>4 weeks | VO<sub>2</sub>peak,<br>gait speed |
| Boyne,<br>2023 | USA | E: 27<br>C: 28 | E: 2.7<br>C: 2.2<br>years | E: 63.8<br>C: 61.5 | Overground<br>walking,<br>Treadmill<br>45 min | 35-60%<br>HRR | Overground<br>walking,<br>treadmill<br>45 min | 30 s,<br>> 60%<br>HRR | 30-60 s,<br>Standing<br>or seated<br>rest | 3<br>days/week<br>12 weeks | gait speed |
| Boyne,<br>2025 | USA | E: 27<br>C: 28 | E: 2.5<br>C: 2.1<br>years | E: 63.8<br>C: 61.5 | Overground<br>walking,<br>Treadmill<br>45 min | 40-60%<br>HRR | Walking<br>exercise<br>45 min | 30 s,<br>maximu<br>m safe<br>speed | 30-60 s,<br>passive<br>recovery | 3<br>days/week<br>12 weeks | VO<sub>2</sub>peak,<br>gait speed |
| Do,<br>2024 | Korea | E: 11<br>C: 11 | E: 88<br>C: 81.5<br>months | E: 61.8<br>C: 63.5 | Aerobic,<br>balance,<br>stretching | – | Robot-assisted<br>walking<br>20 min | 3 min,<br>> 14<br>Borg<br>scale | 2 min,<br>10-11<br>Borg<br>scale | E: 3<br>days/week<br>C: 2<br>days/week<br>8 weeks | VO<sub>2</sub>peak,<br>gait speed |
| Jin, 2012 | China | E: 68<br>C: 65 | E: 18.5<br>C: 17.9<br>months | E: 57<br>C: 56 | Overground<br>walking<br>40 min | 20-30%<br>HRR | Cycle<br>ergometer<br>40 min | 6-10 min,<br>70%<br>HRR | 2-3 min,<br>rest | 5<br>days/week<br>8 weeks | VO<sub>2</sub>max,<br>gait speed |
| Kim 2025 | Korea | E: 17<br>C: 17 | E: 76.1<br>C: 74.1<br>days | E: 70.5<br>C: 70.8 | Treadmill<br>20 min | 40-50%<br>HRR | Skating<br>exercise<br>20 min | 1 min,<br>80%<br>HRR | 1 min,<br>50%<br>HRR | 4<br>days/week<br>12 weeks | VO<sub>2</sub>peak |
| Marzolini,<br>2023 | Canada | E: 24<br>C: 23 | E: 10.8<br>C: 10.6<br>months | E: 62.8<br>C: 60.9 | Treadmill<br>60 min | 60-80%<br>HRR | Treadmill<br>30-32 min | First 4 weeks |  | 5<br>days/week<br>20 weeks | VO<sub>2</sub>peak,<br>gait speed |
|  |  |  |  |  |  |  |  | 30 s,<br>Maximal<br>safe<br>intensity | 60 s,<br>30-40%<br>of peak<br>HR |  |  |
|  |  |  |  |  |  |  |  | After 4 weeks |  |  |  |
|  |  |  |  |  |  |  |  | 2 min | 3 min |  |  |
| Moncion,<br>2024 | Canada | E: 40<br>C: 42 | E: 1.9<br>C: 1.7<br>years | E: 65.4<br>C: 64.4 | Recumbent<br>stepper<br>20 min | 40-60%<br>HRR | Recumbent<br>stepper<br>19 min | 10 min,<br>80%<br>HRR | 9 min,<br>30%<br>HRR | 3<br>days/week,<br>12 weeks | VO<sub>2</sub>peak,<br>gait speed |
| Munari,<br>2018 | Italy | E: 8<br>C: 7 | E: 5.2<br>C: 6.4<br>years | E: 61<br>C: 62 | Treadmill<br>55 min | 60%<br>VO<sub>2</sub>peak | Uphill<br>walking<br>50 min | 5 min,<br>85-95%<br>VO<sub>2</sub>peak | 3 min,<br>50%<br>VO<sub>2</sub>peak | 3<br>days/week<br>12 weeks | VO<sub>2</sub>peak,<br>gait speed |
| Sanberg,<br>2016 | Sweden | E: 29<br>C: 27 | E: 4.9<br>C: 6.3<br>days | E: 71.3<br>C: 70.4 | Standard<br>care | – | Cycle<br>ergometer<br>60 min | 8 min,<br>80%<br>maximal<br>HR | 10 min,<br>low<br>intensity<br>exercise | 3-7<br>days/week,<br>12 weeks | gait speed |
| Soh, 2020 | Korea | E: 18<br>C: 18 | E: 6.5<br>C: 5.9<br>months | E: 56.3<br>C: 57.4 | Treadmill<br>30 min | 50-80%<br>HRR | Skating<br>exercise<br>30 min | 10<br>repetition<br>s,<br>< 14<br>Borg<br>scale | 1 min,<br>stretching | 3<br>days/week<br>12 weeks | VO<sub>2</sub>peak, |

HRR indicates heart rate reserve; VO<sub>2</sub>peak, peak oxygen uptake; “—” indicates that the intensity was not specified or not applicable for the control group.

### Characteristics of Included Studies

All included studies were RCTs published from 2010 onwards, with 7 studies published after 2020.^16,21,25,26,28,29,32^ The studies were conducted across six countries, predominantly in North America [United States (n=3)^16,24,25^ and Canada (n=2)^21,29^]. Common inclusion criteria were as follows: more than 6 months post-stroke in 6 studies;^16,21,24,25,27,30^ stable cardiovascular conditions in 8 studies;^16,21,24,25,28–30,32^ the ability to walk 5–10 meters independently with or without assistive devices in 6 studies;^16,21,24,25,27,31^ the ability to communicate with investigators in 7 studies;^16,21,24,25,29,30,32^ the absence of serious musculoskeletal problems or pain in 8 studies;^16,21,24–26,28,29,32^ and the absence of major poststroke depression in 4 studies.^16,25–27^

### Participants

The 11 studies included in this meta-analysis reported data obtained from 551 participants, of whom 359 (65.2%) were male. The mean age ranged from 56 (SD 7) to 71.3 (SD 7) years, with most studies reporting participants in their 60s. The mean duration since stroke ranged from 4.9 (SD 5.8) days to 6.4 (SD 3.76) years. Three studies included participants within 6 months post-stroke,^28,31,32^ whereas 8 studies included those more than 6 months post-stroke.^16,21,24-^_27,29,30_

### Interventions

Treadmill training was the most common intervention (5 studies),^16,24–26,29^ followed by a cycle ergometer (2 studies)^27,31^ and other modes such as skating exercise, uphill walking, or a recumbent stepper (4 studies).^21,28,30,32^ Exercise intensity was prescribed using the HRR (5 studies),^16,21,27,28,31^ the VO_2_peak (1 study),^30^ the Borg scale (2 studies),^26,32^ or maximal safe criteria (3 studies).^24,25,29^ Three studies included participants younger than 60 years,^24,27,32^ and eight studies included participants aged 60 years or older.^16,21,25,26,28–31^ Three studies were conducted in the subacute stage,^28,31,32^ and eight studies were conducted in the chronic stage.^16,21,24–27,29,30^ Stroke severity was reported to be mild in six studies^21,26,28,30–32^ and moderate in five studies.^16,24,25,27,29^ The intervention time per session exceeded 20 minutes in eight studies^16,24,25,27,29–32^ and was 20 minutes or less in three studies.^21,26,28^ The training frequency was more than three sessions per week in four studies^27–29,31^ and three or fewer sessions in seven studies.^16,21,24–26,30,32^ The intervention length exceeded 8 weeks in eight studies^16,21,25,28–32^ and was 8 weeks or less in three studies.^24,26,27^ For the control groups, the most frequent intervention was low- to moderate-intensity continuous training (9 studies),^16,21,24,25,27–30,32^ and two studies used usual care for the purpose of comparison.^26,31^ Information about the interventions is summarized in Table 3.

**Table 3.**
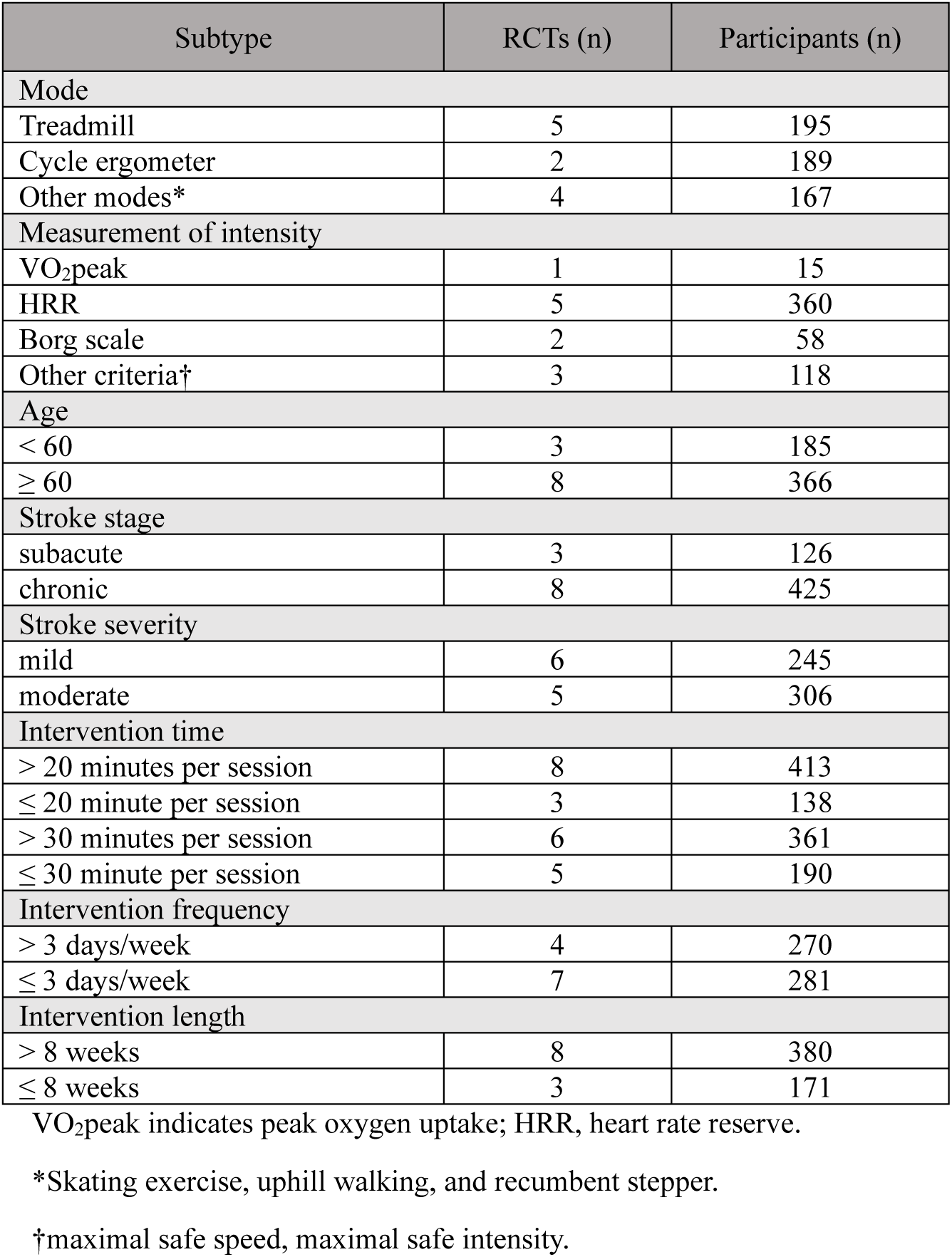
Information Regarding High-Intensity Interval Training in the Included Studies.

### Risk of Bias

No trials were excluded because the PEDro score was less than 4. All the studies reported random allocation, and allocation concealment was applied in nine studies.^16,21,24–26,28–32^ Although the blinding of therapists or participants was not employed, ten studies reported the blinding of outcome assessors for at least one outcome.^16,21,24–30,32^ Seven studies adopted the intention-to-treat principle.^16,21,24–26,29,31^ Baseline characteristics were generally comparable across groups, with only one study reporting a significant baseline imbalance.^24^ Furthermore, all the studies provided point estimates and measures of variability for at least one outcome, and ten of the eleven studies reported data for more than 85% of the initially allocated participants.^16,21,24–31^ On the basis of these criteria, all included RCTs were rated as having high methodological quality. The detailed PEDro assessment results for each study are presented in the Supplemental Material.

### Outcomes

#### Cardiorespiratory Fitness

Nine of the eleven studies assessed VO_2_peak as an outcome. Of these, six reported medium to large effect sizes in favor of HIIT, with the SMD ranging from 0.59 to 2.91. The pooled analysis demonstrated a significant overall improvement in the experimental group, accompanied by considerable between-study heterogeneity compared with the control group (n=440; SMD=1.05 [95% CI, 0.48–1.62]; *P*<0.01; I^2^=85%; MD=3.12 mL/kg/min) (Figure 2a). Funnel plots revealed no apparent asymmetry (Figure 2b), and Egger’s test did not suggest publication bias (*P*=0.95).

**Figure 2.**
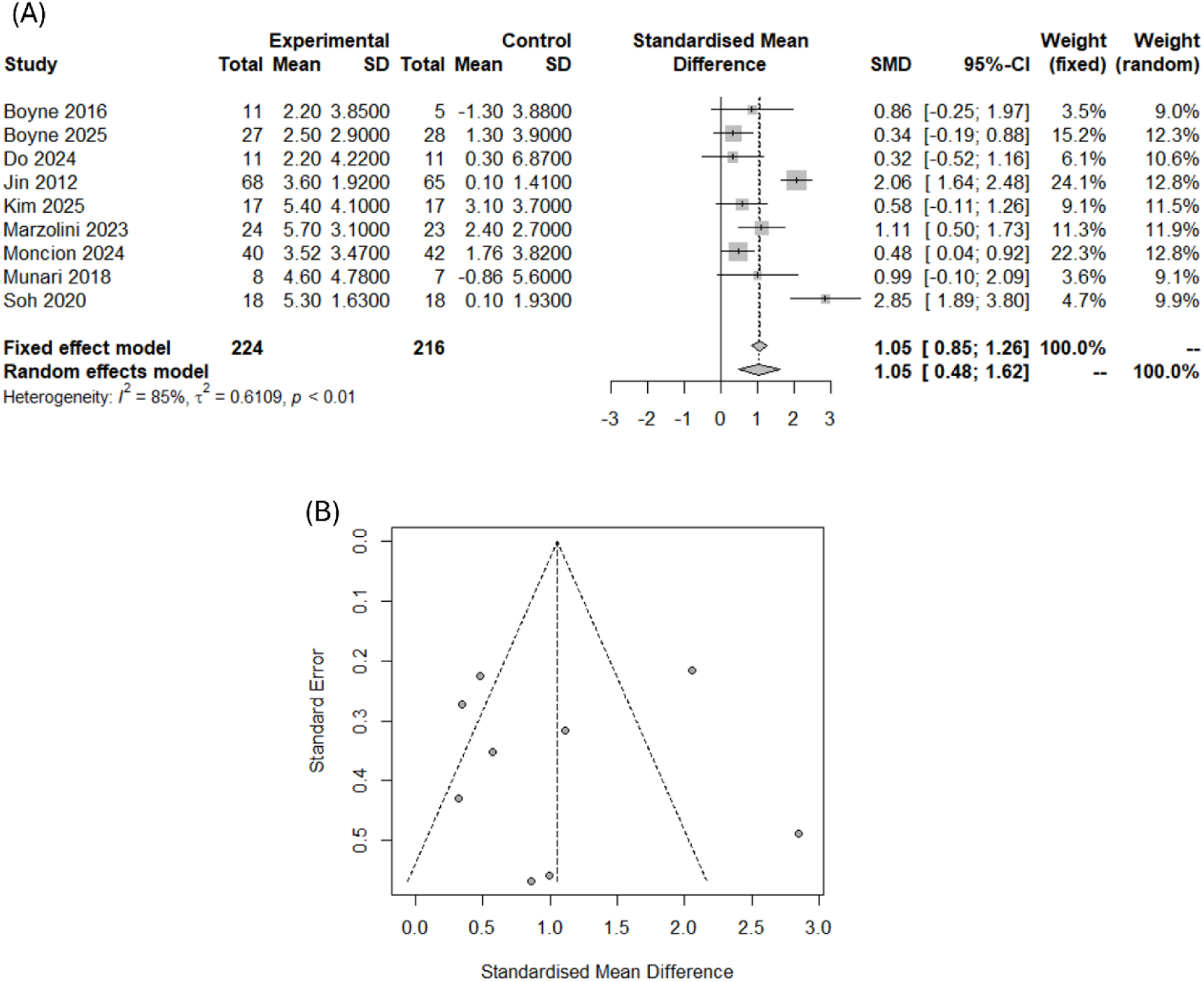
**Meta-Analysis of the Overall Effect Size of HIIT on the Peak Level of Oxygen Uptake (VO_2_peak). (**A) Forest plot of standardized mean differences between the HIIT intervention and control intervention groups. Compared with the control intervention, the HIIT intervention resulted in greater improvements in cardiorespiratory fitness. (B) Funnel plot for publication bias. The funnel plot demonstrates a symmetrical distribution of studies, and no publication bias was detected.

Subgroup analyses revealed significant between-group differences by age and intervention time. Participants aged ≤60 years (3 studies, SMD=1.97, *P*<0.01; MD=4.18 mL/kg/min) achieved greater improvements in VO_2_peak than those aged >60 years did (6 studies, SMD=0.58, *P*<0.01; MD=2.23 mL/kg/min), and intervention sessions that exceeded 20 minutes (6 studies, SMD=1.37, *P*<0.01; MD=3.54 mL/kg/min) were more effective than intervention sessions that were 20 minutes or less (3 studies, SMD=0.48, *P*<0.01; MD=1.9 mL/kg/min). In contrast, no significant subgroup differences were observed for stroke stage, stroke severity, intervention frequency, or intervention length. Sensitivity analysis using the Copas selection model confirmed that the pooled estimate for VO_2_peak was robust to potential publication bias. The results remained consistent when the random-effects model was compared with the Copas-adjusted model, and a visual inspection of the funnel and contour plots did not suggest substantial asymmetry (Supplemental Material).

#### Walking Ability

Nine of the eleven studies examined gait speed; among these studies, five reported medium to large effect sizes ranging from 0.53 to 1.61. The pooled analysis indicated a small but significant overall effect of HIIT on gait speed compared with the control group, although between-study heterogeneity was high (n=481; SMD=0.61 [95% CI, 0.19–1.02]; *P*<0.01; I^2^=77%; n=292; MD=0.11 m/s) (Figure 3a). Funnel plots did not indicate asymmetry (Figure 3b), and Egger’s test suggested that there was no publication bias (*P*=0.13).

**Figure 3.**
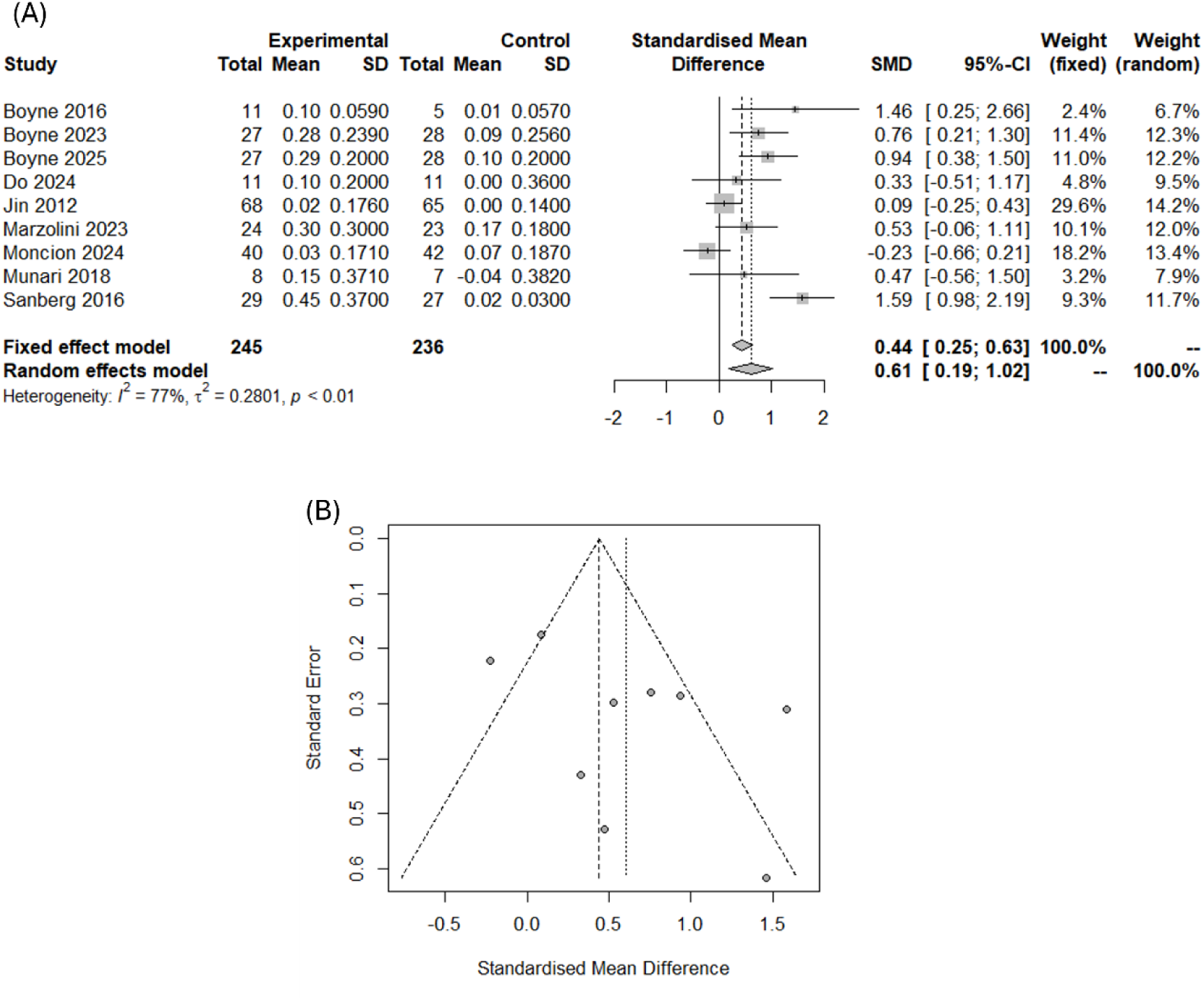
**Meta-analysis of the overall effect size of HIIT on gait speed. (**A) Forest plot of standardized mean differences between the HIIT intervention and control intervention groups. HIIT resulted in greater improvements in walking ability than the control intervention did. (B) Funnel plot for publication bias. The funnel plot demonstrates a symmetrical distribution of studies, and no publication bias was detected.

**Figure 4.**
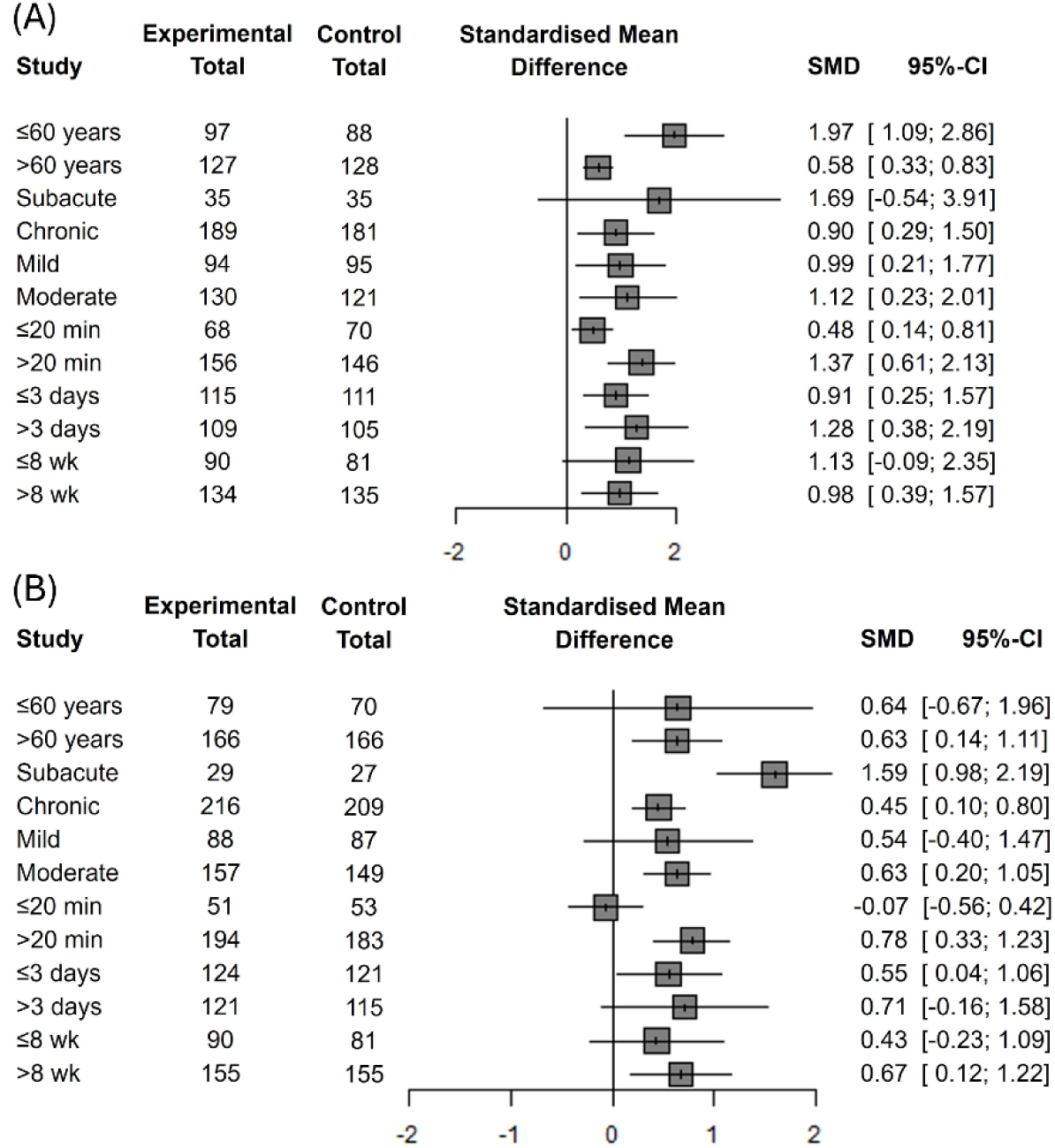
Subgroup Analysis of HIIT on Cardiorespiratory fitness and Walking Ability. Subgroup analysis in terms of participant characteristics (age, time since stroke, and stroke severity) and intervention dose (session time, frequency, and program duration). (A) Subgroup analysis for cardiorespiratory fitness. The effects of HIIT were significantly greater in participants under 60 years and in those with training sessions lasting more than 20 minutes. (B) Subgroup analysis for walking ability. HIIT showed a significantly greater effects on participants in the subacute phase and in those with training sessions lasting more than 20 minutes.

Subgroup analyses revealed significant between-group differences according to stroke stage and intervention time. Participants in the subacute stage (1 study, SMD=1.59, *P*<0.01) exhibited greater gains in gait speed than those in the chronic stage did (8 studies, SMD=0.45, *P*<0.05), and intervention sessions longer than 20 minutes (7 studies, SMD=0.78, *P*<0.01; MD=0.13 m/s) were superior to those lasting 20 minutes or less (2 studies, SMD=-0.07, *P*=0.78; MD=-0.02 m/s). No significant subgroup differences were observed in terms of age, stroke severity, intervention frequency, or intervention length. Sensitivity analysis using the Copas selection model confirmed the robustness of the pooled estimate, as the adjusted results were consistent with those of the random-effects model, and visual inspection of the funnel and contour plots did not reveal substantial asymmetry (Supplemental Material).

### Adverse Events

Overall, 9 of the 11 studies reported on the safety of HIIT.^16,21,24,26,28–32^ The remaining two studies did not provide information on adverse events.^25,27^ Among these nine studies, five reported no adverse events,^26,28,30–32^ and four reported no serious adverse events.^16,21,24,29^ Only three RCTs provided detailed descriptions of treatment-related adverse events,^16,24,29^ which included musculoskeletal complaints (joint or muscle pain), fatigue, light-headedness, and nausea. Two studies reported adverse events that occurred during the experimental period, namely, a fall and a recurrent stroke, both of which were detemined to be unrelated to the intervention.^16,29^

#### Certainty of Evidence

With respect to CRF, a high degree of statistical heterogeneity (I^2^=85%) indicated substantial inconsistency, which led to a decrease in the certainty of evidence. Similarly, the evidence for walking ability presented substantial inconsistency (I^2^=77%), also resulting in a downgrade. Consequently, according to the GRADE approach, the final certainty for both outcomes was rated as moderate (Table 4).

**Table 4.** Grading of Recommendations, Assessment, Development and Evaluation (GRADE) Summary of Findings of rtainty of Evidence.

| Quality assessments |  |  |  |  |  |  | Number of patients | Effect | Quality |
| --- | --- | --- | --- | --- | --- | --- | --- | --- | --- |
| Number of studies | Study design | Risk of bias | Inconsistency | Indirectness | Imprecision | Other considerations | HIIT | SMD (95% CI) |  |
| 9 | RCT | Not serious | Serious* | Not serious | Not serious | None | 224/440 | 1.05 (0.48-1.62) | ⊕⊕⊕○<br>Moderate |
| 9 | RCT | Not serious | Serious* | Not serious | Not serious | None | 245/481 | 0.61 (0.19-1.02) | ⊕⊕⊕○<br>Moderate |

HIIT indicates high-intensity interval training; SMD, standardized mean difference; CI, confidence interval; VO2peak, peak oxygen uptake. High quality: very confident that the true effect lies close to that of the estimate of the effect; Moderate quality: the true effect is likely to be close to the estimate of the effect, but there is a possibility that it is substantially different; Low quality: the true effect may be substantially different from the estimate of the effect; and Very low quality: the true effect is likely to be substantially different from the estimate of the effect.
\*Actual therapeutic benefits may differ substantially across patient population or intervention settings.

## Discussion

The present systematic review and meta-analysis evaluated the effectiveness of HIIT compared with that of low- to moderate-intensity training or conventional rehabilitation programs for improving CRF and walking ability in patients with stroke. Eleven high-quality RCTs involving 551 poststroke participants were included. Our findings demonstrate that HIIT provides significant benefits for VO_2_peak and gait speed, both of which are critical indicators of cardiometabolic health and mobility function. Importantly, no serious adverse events related to exercise were reported, thus suggesting that HIIT can be safely implemented for patients with stroke when it is performed under appropriate supervision.

Compared with nonaerobic interventions, aerobic exercise in stroke rehabilitation has been shown to improve CRF by enhancing aerobic capacity and walking economy.^33^ Higher exercise intensity appears to be particularly effective at improving peripheral muscular oxidative capacity and aerobic performance.^34,35^ While a recent systematic review suggested that HIIT may improve only certain aspects of CRF (e.g., VT1) but not VO_2_peak,^36^ our analysis provides stronger evidence indicating that HIIT significantly enhances CRF in stroke survivors. We observed moderate-to-large effect sizes, with a mean improvement in the VO_2_peak of 3.12 mL/kg/min (95% CI, 2.15–4.09). These results are consistent with the HIIT-usual care difference of 2.89 mL/kg/min improvement and greater than the HIIT-MICT difference of 0.82 mL/kg/min improvement reported in a recent network meta-analysis study.^37^

Using the GRADE framework, we are moderately confident that HIIT is among the most effective interventions for improving VO_2_peak. This moderate certainty rating is due to inconsistency across studies, which indicates that the true effect might be different. Despite this inconsistency, the observed increase in VO_2_peak of more than 1.0 mL/kg/ with HIIT is clinically meaningful. An increase of 1.0 mL/kg/min in VO_2_peak is associated with a 15% reduction in cardiovascular mortality,^38^ whereas a 1-MET increase (3.0 mL/kg/min in stroke) is associated with a 7% reduction in stroke hospitalization and a 9% lower risk of ischemic stroke.^3,39^ This improvement in CRF appears to be driven by adaptations in both central cardiac function and peripheral skeletal muscle utilization. Previous research has suggested that compared with MICT, HIIT leads to greater improvements in stroke volume (central) while enhancing skeletal muscle oxidative capacity by reversing mitochondrial dysfunction and improving calcium handling to delay fatigue (peripheral).^40^

Subgroup analysis revealed a more pronounced improvement in VO_2_peak (4.18 mL/kg/min) among middle-aged participants (≤ 60 years). This improvement may be attributable to a superior motor learning capacity and greater neuroplastic potential in younger individuals.^41^

Furthermore, when VO_2_peak levels are comparably reduced, younger individuals may achieve greater improvements, as the magnitude of reduction relative to normative values is greater in this population.^42^ No significant subgroup differences were observed for stroke stage or severity, indicating that HIIT is effective not only for patients with mild impairments but also for those with moderate impairments and that sufficient improvements in CRF can be achieved in both the subacute and chronic phases.

With respect to intervention dose, greater improvements in VO_2_peak were observed when the duration of each session exceeded 20 minutes, whereas increasing the frequency (>3 days per week) or overall intervention length (>8 weeks) did not yield additional benefits. While aerobic exercise generally produces greater improvements in CRF with higher training doses, delivering such intensity programs to stroke survivors poses practical challenges. A recent review revealed that the majority of high-intensity training programs require more than 30 minutes per session over 12 weeks to improve CRF.^14^ In contrast, our findings suggest that a HIIT protocol of just 20–30 minutes per session, three times a week for eight weeks, is also effective for enhancing CRF in stroke survivors. This finding highlights the fact that HIIT can be a particularly time-efficient strategy for improving CRF in stroke patients.

The results of the present meta-analysis support the notion that compared with conventional rehabilitation, high-intensity aerobic exercise, including low- to moderate-intensity aerobic exercise, can improve walking ability. Walking ability is a key measure of mobility and an important predictor of family and community ambulation.^43^ The certainty of this estimate is moderate, reflecting the high level of heterogeneity. Improvements in gait speed may contribute to increased participation and activities of daily living among stroke survivors.^44^ Evidence indicates that gains in CRF following HIIT lower the energy cost of walking and improve endurance,^30^ which translates to a faster and more efficient gait. Given that stroke patients have a 1.5- to 2.0-fold higher energy cost of walking than healthy individuals do, improving CRF is essential for restoring their ability to walk.^45^ From a neurophysiological perspective, potential mechanisms include increased corticospinal excitability and neurotrophin expression, which facilitate motor learning,^46,47^ coupled with the promotion of neuroplasticity and neuroprotection.^48,49^ The recovery intervals in HIIT could also provide opportunities for mental rehearsal and cognitive processing beyond those afforded by conventional gait training, potentially leading to better retention of motor learning.^50^

The subgroup analysis results suggested that time since stroke onset may be an important factor influencing the effectiveness of HIIT on walking ability in one subgroup study of the subacute phase. Similar to VO_2_peak, intervention sessions with a duration exceeding 20 minutes resulted in greater improvements in gait speed. Therefore, although HIIT offers the advantage of efficiency within shorter exercise durations because of its interval-based structure, our findings suggest that a minimum intervention time of approximately 20 minutes may be required to achieve meaningful improvements in both CRF and walking ability.

When a HIIT protocol is implemented for stroke rehabilitation, clinicians may be concerned about actual or potential safety risks. Two studies reported adverse events (i.e., a fall and recurrent stroke), neither of which was directly attributable to HIIT.^16,29^ Other side effects were minor and within the expected range for older individuals engaging in exercise, for example, fatigue and musculoskeletal pain. These findings may reflect both the inherent safety of HIIT and the relatively high functional status of participants, which was influenced by the inclusion and exclusion criteria of the trials. Furthermore, safety analyses indicated that poststroke HIIT was not associated with cardiovascular complications (e.g., hypotension, hypertension, and arrhythmia) or orthopedic events (e.g., pain and falls).^51^

To ensure safety in the process applying HIIT after stroke, baseline medical screening and, when possible, graded exercise stress testing are recommended.^52^ Individuals with high-risk cardiovascular or orthopedic conditions should be excluded, and both objective indicators (e.g., heart rate and blood pressure) and subjective measures (e.g., perceived exertion) should be monitored throughout training.^17^ The use of safety equipment, such as harnesses or orthotic devices, can further reduce the risk of falls or injuries.^16^ With these precautions, HIIT can be implemented safely and effectively in poststroke rehabilitation.

There are several limitations to acknowledge. First, the primary limitation of this meta-analysis is the substantial between-study heterogeneity observed for both primary outcomes, i.e., CRF (I2=85%) and walking ability (I2=77%). This inconsistency is likely attributable to the variability across the included trials in terms of HIIT protocols (e.g., exercise session duration and intensity) and the diverse characteristics of the patient populations, including age, time since stroke and severity of impairment. Consequently, while the pooled estimates indicate a significant overall benefit, they should be interpreted with caution, as the true treatment effect may vary substantially depending on the specific clinical context and intervention environment. Second, while we conducted several subgroup analyses to explore these sources of heterogeneity, their statistical power was limited by the small number of studies within certain categories. Future research incorporating larger trial numbers within specific subgroups is needed to draw more definitive conclusions about the optimal HIIT protocols for distinct stroke populations.

## Conclusion

This systematic review and meta-analysis demonstrated that HIIT significantly improved CRF and walking ability among stroke survivors, with moderate-certainty evidence according to the GRADE framework. These findings indicate that HIIT is an effective, safe, and time-efficient rehabilitation strategy that may complement or surpass conventional exercise programs in poststroke rehabilitation. Greater improvements in CRF were observed among middle-aged participants, while the benefits were consistent regardless of stroke severity or onset time. The magnitude of improvement exceeded clinically meaningful thresholds and was particularly evident when the session duration exceeded 20 minutes. Moreover, interventions delivered three times per week for approximately eight weeks were sufficient to produce meaningful gains in CRF and walking ability.

## Data Availability

This study is a meta-analysis and uses data extracted from previously published studies, all of which are publicly available in the original articles. No new datasets were generated or analyzed during the current study.

## Acknowledgments

None.

## Abbreviations and Acronyms

CRF: Cardiorespiratory fitness
HIIT: High-intensity interval training
HRR: Heart rate reserve
MD: Mean difference
MICT: Moderate-intensity continuous training
RPE: Ratings of Perceived Exertion
SMD: Standardized mean difference
VO2peak: Peak oxygen uptake

## References

1. Group GBDNDC. Global, regional, and national burden of neurological disorders during 1990-2015: a systematic analysis for the Global Burden of Disease Study 2015. Lancet Neurol. 2017;16:877–897. doi: 10.1016/S1474-4422(17)30299-5

2. Collaborators GBDS. Global, regional, and national burden of stroke and its risk factors, 1990-2019: a systematic analysis for the Global Burden of Disease Study 2019. Lancet Neurol. 2021;20:795–820. doi: 10.1016/S1474-4422(21)00252-0

3. Ehrman JK, Keteyian SJ, Johansen MC, Blaha MJ, Al-Mallah MH, Brawner CA. Improved cardiorespiratory fitness is associated with lower incident ischemic stroke risk: Henry Ford FIT project. J Stroke Cerebrovasc Dis. 2023;32:107240. doi: 10.1016/j.jstrokecerebrovasdis.2023.107240

4. Prestgaard E, Mariampillai J, Engeseth K, Erikssen J, Bodegard J, Liestol K, Gjesdal K, Kjeldsen S, Grundvold I, Berge E. Change in Cardiorespiratory Fitness and Risk of Stroke and Death: Long-Term Follow-Up of Healthy Middle-Aged Men. Stroke. 2019;50:155–161. doi: 10.1161/STROKEAHA.118.021798

5. Kelly JO, Kilbreath SL, Davis GM, Zeman B, Raymond J. Cardiorespiratory fitness and walking ability in subacute stroke patients. Arch Phys Med Rehabil. 2003;84:1780– 1785. doi: 10.1016/s0003-9993(03)00376-9

6. Billinger SA, Coughenour E, Mackay-Lyons MJ, Ivey FM. Reduced cardiorespiratory fitness after stroke: biological consequences and exercise-induced adaptations. Stroke Res Treat. 2012;2012:959120. doi: 10.1155/2012/959120

7. Ivey FM, Hafer-Macko CE, Macko RF. Task-oriented treadmill exercise training in chronic hemiparetic stroke. J Rehabil Res Dev. 2008;45:249–259. doi: 10.1682/jrrd.2007.02.0035

8. Pase MP, Beiser A, Enserro D, Xanthakis V, Aparicio H, Satizabal CL, Himali JJ, Kase CS, Vasan RS, DeCarli C, et al. Association of Ideal Cardiovascular Health With Vascular Brain Injury and Incident Dementia. Stroke. 2016;47:1201–1206. doi: 10.1161/STROKEAHA.115.012608

9. Smith AC, Saunders DH, Mead G. Cardiorespiratory fitness after stroke: a systematic review. Int J Stroke. 2012;7:499–510. doi: 10.1111/j.1747-4949.2012.00791.x

10. Jorgensen HS, Nakayama H, Raaschou HO, Olsen TS. Recovery of walking function in stroke patients: the Copenhagen Stroke Study. Arch Phys Med Rehabil. 1995;76:27–32. doi: 10.1016/s0003-9993(95)80038-7

11. Hill K, Ellis P, Bernhardt J, Maggs P, Hull S. Balance and mobility outcomes for stroke patients: a comprehensive audit. Aust J Physiother. 1997;43:173–180. doi: 10.1016/s0004-9514(14)60408-6

12. MacKay-Lyons M, Billinger SA, Eng JJ, Dromerick A, Giacomantonio N, Hafer-Macko C, Macko R, Nguyen E, Prior P, Suskin N, et al. Aerobic Exercise Recommendations to Optimize Best Practices in Care After Stroke: AEROBICS 2019 Update. Phys Ther. 2020;100:149–156. doi: 10.1093/ptj/pzz153

13. Balady GJ, Williams MA, Ades PA, Bittner V, Comoss P, Foody JM, Franklin B, Sanderson B, Southard D, American Heart Association Exercise CR, et al. Core components of cardiac rehabilitation/secondary prevention programs: 2007 update: a scientific statement from the American Heart Association Exercise, Cardiac Rehabilitation, and Prevention Committee, the Council on Clinical Cardiology; the Councils on Cardiovascular Nursing, Epidemiology and Prevention, and Nutrition, Physical Activity, and Metabolism; and the American Association of Cardiovascular and Pulmonary Rehabilitation. Circulation. 2007;115:2675–2682. doi: 10.1161/CIRCULATIONAHA.106.180945

14. Luo L, Meng H, Wang Z, Zhu S, Yuan S, Wang Y, Wang Q. Effect of high-intensity exercise on cardiorespiratory fitness in stroke survivors: A systematic review and meta-analysis. Ann Phys Rehabil Med. 2020;63:59–68. doi: 10.1016/j.rehab.2019.07.006

15. Baricich A, Borg MB, Battaglia M, Facciorusso S, Spina S, Invernizzi M, Scotti L, Cosenza L, Picelli A, Santamato A. High-Intensity Exercise Training Impact on Cardiorespiratory Fitness, Gait Ability, and Balance in Stroke Survivors: A Systematic Review and Meta-Analysis. J Clin Med. 2024;13. doi: 10.3390/jcm13185498

16. Boyne P, Billinger SA, Reisman DS, Awosika OO, Buckley S, Burson J, Carl D, DeLange M, Doren S, Earnest M, et al. Optimal Intensity and Duration of Walking Rehabilitation in Patients With Chronic Stroke: A Randomized Clinical Trial. JAMA Neurol. 2023;80:342–351. doi: 10.1001/jamaneurol.2023.0033

17. Boyne P, Billinger S, MacKay-Lyons M, Barney B, Khoury J, Dunning K. Aerobic Exercise Prescription in Stroke Rehabilitation: A Web-Based Survey of US Physical Therapists. J Neurol Phys Ther. 2017;41:119–128. doi: 10.1097/NPT.0000000000000177

18. Oliveira A, Fidalgo A, Farinatti P, Monteiro W. Effects of high-intensity interval and continuous moderate aerobic training on fitness and health markers of older adults: A systematic review and meta-analysis. Arch Gerontol Geriatr. 2024;124:105451. doi: 10.1016/j.archger.2024.105451

19. Sert H, Gulbahar Eren M, Gurcay B, Koc F. The effectiveness of a high-intensity interval exercise on cardiometabolic health and quality of life in older adults: a systematic review and meta-analysis. BMC Sports Sci Med Rehabil. 2025;17:128. doi: 10.1186/s13102-025-01176-5

20. Dlugosz EM, Chappell MA, Meek TH, Szafranska PA, Zub K, Konarzewski M, Jones JH, Bicudo JE, Nespolo RF, Careau V, et al. Phylogenetic analysis of mammalian maximal oxygen consumption during exercise. J Exp Biol. 2013;216:4712–4721. doi: 10.1242/jeb.088914

21. Moncion K, Rodrigues L, De Las Heras B, Noguchi KS, Wiley E, Eng JJ, MacKay-Lyons M, Sweet SN, Thiel A, Fung J, et al. Cardiorespiratory Fitness Benefits of High-Intensity Interval Training After Stroke: A Randomized Controlled Trial. Stroke. 2024;55:2202–2211. doi: 10.1161/STROKEAHA.124.046564

22. Schmid A, Duncan PW, Studenski S, Lai SM, Richards L, Perera S, Wu SS. Improvements in speed-based gait classifications are meaningful. Stroke. 2007;38:2096–2100. doi: 10.1161/STROKEAHA.106.475921

23. Guyatt GH, Oxman AD, Vist GE, Kunz R, Falck-Ytter Y, Alonso-Coello P, Schunemann HJ, Group GW. GRADE: an emerging consensus on rating quality of evidence and strength of recommendations. BMJ. 2008;336:924–926. doi: 10.1136/bmj.39489.470347.AD

24. Boyne P, Dunning K, Carl D, Gerson M, Khoury J, Rockwell B, Keeton G, Westover J, Williams A, McCarthy M, et al. High-Intensity Interval Training and Moderate-Intensity Continuous Training in Ambulatory Chronic Stroke: Feasibility Study. Phys Ther. 2016;96:1533–1544. doi: 10.2522/ptj.20150277

25. Boyne P, Miller A, Schwab-Farrell SM, Sucharew H, Carl D, Billinger SA, Reisman DS. Training Parameters and Adaptations That Mediate Walking Capacity Gains from High-Intensity Gait Training Poststroke. Med Sci Sports Exerc. 2025;57:1285–1296. doi: 10.1249/MSS.0000000000003691

26. Do J, Lim WT, Kim DY, Ko EJ, Ko MH, Kim GW, Kim JH, Kim S, Kim H. Effects of high-intensity interval robot-assisted gait training on cardiopulmonary function and walking ability in chronic stroke survivors: A multicenter single-blind randomized controlled trial. J Back Musculoskelet Rehabil. 2024;37:1309–1319. doi: 10.3233/BMR-230385

27. Jin H, Jiang Y, Wei Q, Wang B, Ma G. Intensive aerobic cycling training with lower limb weights in Chinese patients with chronic stroke: discordance between improved cardiovascular fitness and walking ability. Disabil Rehabil. 2012;34:1665–1671. doi: 10.3109/09638288.2012.658952

28. Kim MS. The Effect of Skating Exercises as High-Intensity Interval Training on Elderly Stroke Patients. Brain Sci. 2025;15. doi: 10.3390/brainsci15070676

29. Marzolini S, Robertson AD, MacIntosh BJ, Corbett D, Anderson ND, Brooks D, Koblinsky N, Oh P. Effect of High-Intensity Interval Training and Moderate-Intensity Continuous Training in People With Poststroke Gait Dysfunction: A Randomized Clinical Trial. J Am Heart Assoc. 2023;12:e031532. doi: 10.1161/JAHA.123.031532

30. Munari D, Pedrinolla A, Smania N, Picelli A, Gandolfi M, Saltuari L, Schena F. High-intensity treadmill training improves gait ability, VO2peak and cost of walking in stroke survivors: preliminary results of a pilot randomized controlled trial. Eur J Phys Rehabil Med. 2018;54:408–418. doi: 10.23736/S1973-9087.16.04224-6

31. Sandberg K, Kleist M, Falk L, Enthoven P. Effects of Twice-Weekly Intense Aerobic Exercise in Early Subacute Stroke: A Randomized Controlled Trial. Arch Phys Med Rehabil. 2016;97:1244–1253. doi: 10.1016/j.apmr.2016.01.030

32. Soh SH, Joo MC, Yun NR, Kim MS. Randomized Controlled Trial of the Lateral Push-Off Skater Exercise for High-Intensity Interval Training vs Conventional Treadmill Training. Arch Phys Med Rehabil. 2020;101:187–195. doi: 10.1016/j.apmr.2019.08.480

33. Saunders DH, Sanderson M, Hayes S, Kilrane M, Greig CA, Brazzelli M, Mead GE. Physical fitness training for stroke patients. Cochrane Database Syst Rev. 2016;3:CD003316. doi: 10.1002/14651858.CD003316.pub6

34. Egan B, Carson BP, Garcia-Roves PM, Chibalin AV, Sarsfield FM, Barron N, McCaffrey N, Moyna NM, Zierath JR, O’Gorman DJ. Exercise intensity-dependent regulation of peroxisome proliferator-activated receptor coactivator-1 mRNA abundance is associated with differential activation of upstream signalling kinases in human skeletal muscle. J Physiol. 2010;588:1779–1790. doi: 10.1113/jphysiol.2010.188011

35. Haskell WL, Lee IM, Pate RR, Powell KE, Blair SN, Franklin BA, Macera CA, Heath GW, Thompson PD, Bauman A. Physical activity and public health: updated recommendation for adults from the American College of Sports Medicine and the American Heart Association. Med Sci Sports Exerc. 2007;39:1423–1434. doi: 10.1249/mss.0b013e3180616b27

36. Wiener J, McIntyre A, Janssen S, Chow JT, Batey C, Teasell R. Effectiveness of High-Intensity Interval Training for Fitness and Mobility Post Stroke: A Systematic Review. PM R. 2019;11:868–878. doi: 10.1002/pmrj.12154

37. Moncion K, Rodrigues L, Wiley E, Noguchi KS, Negm A, Richardson J, MacDonald MJ, Roig M, Tang A. Aerobic exercise interventions for promoting cardiovascular health and mobility after stroke: a systematic review with Bayesian network meta-analysis. Br J Sports Med. 2024;58:392–400. doi: 10.1136/bjsports-2023-107956

38. Keteyian SJ, Brawner CA, Savage PD, Ehrman JK, Schairer J, Divine G, Aldred H, Ophaug K, Ades PA. Peak aerobic capacity predicts prognosis in patients with coronary heart disease. Am Heart J. 2008;156:292–300. doi: 10.1016/j.ahj.2008.03.017

39. Pandey A, Patel MR, Willis B, Gao A, Leonard D, Das SR, Defina L, Berry JD. Association Between Midlife Cardiorespiratory Fitness and Risk of Stroke: The Cooper Center Longitudinal Study. Stroke. 2016;47:1720–1726. doi: 10.1161/STROKEAHA.115.011532

40. Crozier J, Roig M, Eng JJ, MacKay-Lyons M, Fung J, Ploughman M, Bailey DM, Sweet SN, Giacomantonio N, Thiel A, et al. High-Intensity Interval Training After Stroke: An Opportunity to Promote Functional Recovery, Cardiovascular Health, and Neuroplasticity. Neurorehabil Neural Repair. 2018;32:543–556. doi: 10.1177/1545968318766663

41. Lee J, Stone AJ. Combined Aerobic and Resistance Training for Cardiorespiratory Fitness, Muscle Strength, and Walking Capacity after Stroke: A Systematic Review and Meta-Analysis. J Stroke Cerebrovasc Dis. 2020;29:104498. doi: 10.1016/j.jstrokecerebrovasdis.2019.104498

42. Taylor JL, Medina-Inojosa JR, Chacin-Suarez A, Smith JR, Squires RW, Thomas RJ, Johnson BD, Olson TP, Bonikowske AR. Age-Related Differences for Cardiorespiratory Fitness Improvement in Patients Undergoing Cardiac Rehabilitation. Front Cardiovasc Med. 2022;9:872757. doi: 10.3389/fcvm.2022.872757

43. Fulk GD, He Y, Boyne P, Dunning K. Predicting Home and Community Walking Activity Poststroke. Stroke. 2017;48:406–411. doi: 10.1161/STROKEAHA.116.015309

44. Eng JJ, Tang PF. Gait training strategies to optimize walking ability in people with stroke: a synthesis of the evidence. Expert Rev Neurother. 2007;7:1417–1436. doi: 10.1586/14737175.7.10.1417

45. da Cunha IT, Jr., Lim PA, Qureshy H, Henson H, Monga T, Protas EJ. Gait outcomes after acute stroke rehabilitation with supported treadmill ambulation training: a randomized controlled pilot study. Arch Phys Med Rehabil. 2002;83:1258–1265. doi: 10.1053/apmr.2002.34267

46. Mang CS, Snow NJ, Campbell KL, Ross CJ, Boyd LA. A single bout of high-intensity aerobic exercise facilitates response to paired associative stimulation and promotes sequence-specific implicit motor learning. J Appl Physiol (1985). 2014;117:1325–1336. doi: 10.1152/japplphysiol.00498.2014

47. Skriver K, Roig M, Lundbye-Jensen J, Pingel J, Helge JW, Kiens B, Nielsen JB. Acute exercise improves motor memory: exploring potential biomarkers. Neurobiol Learn Mem. 2014;116:46–58. doi: 10.1016/j.nlm.2014.08.004

48. Mang CS, Campbell KL, Ross CJ, Boyd LA. Promoting neuroplasticity for motor rehabilitation after stroke: considering the effects of aerobic exercise and genetic variation on brain-derived neurotrophic factor. Phys Ther. 2013;93:1707–1716. doi: 10.2522/ptj.20130053

49. Austin MW, Ploughman M, Glynn L, Corbett D. Aerobic exercise effects on neuroprotection and brain repair following stroke: a systematic review and perspective. Neurosci Res. 2014;87:8–15. doi: 10.1016/j.neures.2014.06.007

50. Hanlon RE. Motor learning following unilateral stroke. Arch Phys Med Rehabil. 1996;77:811–815. doi: 10.1016/s0003-9993(96)90262-2

51. Carl DL, Boyne P, Rockwell B, Gerson M, Khoury J, Kissela B, Dunning K. Preliminary safety analysis of high-intensity interval training (HIIT) in persons with chronic stroke. Appl Physiol Nutr Metab. 2017;42:311–318. doi: 10.1139/apnm-2016-0369

52. Billinger SA, Arena R, Bernhardt J, Eng JJ, Franklin BA, Johnson CM, MacKay-Lyons M, Macko RF, Mead GE, Roth EJ, et al. Physical activity and exercise recommendations for stroke survivors: a statement for healthcare professionals from the American Heart Association/American Stroke Association. Stroke. 2014;45:2532– 2553. doi: 10.1161/STR.0000000000000022

