## Supplemental Material for "Effects of High-Intensity Interval Training on Cardiorespiratory Fitness in Patients with Stroke: A Meta-Analysis"

**Supplemental Material**
**PRISMA checklist**

| Section and Topic | Item # | Checklist item | Location where item is reported |
|---|---|---|---|
| **TITLE** | | | |
| Title | 1 | Identify the report as a systematic review. | P1 Ln 2-3 |
| **ABSTRACT** | | | |
| Abstract | 2 | See the PRISMA 2020 for Abstracts checklist. | P2 Ln 21-43 |
| **INTRODUCTION** | | | |
| Rationale | 3 | Describe the rationale for the review in the context of existing knowledge. | P5 Ln 89-96 |
| Objectives | 4 | Provide an explicit statement of the objective(s) or question(s) the review addresses. | P5 Ln 97-102 |
| **METHODS** | | | |
| Eligibility criteria | 5 | Specify the inclusion and exclusion criteria for the review and how studies were grouped for the syntheses. | P7 Ln 150-156 |
| Information sources | 6 | Specify all databases, registers, websites, organisations, reference lists and other sources searched or consulted to identify studies. Specify the date when each source was last searched or consulted. | P6 Ln 132-135 |
| Search strategy | 7 | Present the full search strategies for all databases, registers and websites, including any filters and limits used. | P6 Ln 139-140 |
| Selection process | 8 | Specify the methods used to decide whether a study met the inclusion criteria of the review, including how many reviewers screened each record and each report retrieved, whether they worked independently, and if applicable, details of automation tools used in the process. | P7 Ln 141-143 |
| Data collection process | 9 | Specify the methods used to collect data from reports, including how many reviewers collected data from each report, whether they worked independently, any processes for obtaining or confirming data from study investigators, and if applicable, details of automation tools used in the process. | P7 Ln 144-146 |
| Data items | 10a | List and define all outcomes for which data were sought. Specify whether all results that were compatible with each outcome domain in each study were sought (e.g. for all measures, time points, analyses), and if not, the methods used to decide which results to collect. | P6 Ln 117-125 |
| | 10b | List and define all other variables for which data were sought (e.g. participant and intervention characteristics, funding sources). Describe any assumptions made about any missing or unclear information. | P6 Ln 126-129 |
| Study risk of bias assessment | 11 | Specify the methods used to assess risk of bias in the included studies, including details of the tool(s) used, how many reviewers assessed each study and whether they worked independently, and if applicable, details of automation tools used in the process. | P8 Ln 164-168 |
| Effect measures | 12 | Specify for each outcome the effect measure(s) (e.g. risk ratio, mean difference) used in the synthesis or presentation of results. | P9 Ln 198-200 |
| Synthesis methods | 13a | Describe the processes used to decide which studies were eligible for each synthesis (e.g. tabulating the study intervention characteristics and comparing against the planned groups for each synthesis (item #5)). | P8 Ln 176-183 |
| | 13b | Describe any methods required to prepare the data for presentation or synthesis, such as handling of missing summary statistics, or data conversions. | P9 Ln 184-188 |

| Section and Topic | Item # | Checklist item | Location where item is reported |
|---|---|---|---|
| | 13c | Describe any methods used to tabulate or visually display results of individual studies and syntheses. | P10 Ln 204-205 |
| | 13d | Describe any methods used to synthesize results and provide a rationale for the choice(s). If meta-analysis was performed, describe the model(s), method(s) to identify the presence and extent of statistical heterogeneity, and software package(s) used. | P9-10 Ln 196-204 |
| | 13e | Describe any methods used to explore possible causes of heterogeneity among study results (e.g. subgroup analysis, meta-regression). | P10 Ln 206-209 |
| | 13f | Describe any sensitivity analyses conducted to assess robustness of the synthesized results. | P10 Ln 211-216 |
| Reporting bias assessment | 14 | Describe any methods used to assess risk of bias due to missing results in a synthesis (arising from reporting biases). | P10 Ln 210-211 |
| Certainty assessment | 15 | Describe any methods used to assess certainty (or confidence) in the body of evidence for an outcome. | P8 Ln 169-173 |
| **RESULTS** | | | |
| Study selection | 16a | Describe the results of the search and selection process, from the number of records identified in the search to the number of studies included in the review, ideally using a flow diagram. | P10 Ln 220-225 |
| | 16b | Cite studies that might appear to meet the inclusion criteria, but which were excluded, and explain why they were excluded. | P7-8 Ln 157-161 |
| Study characteristics | 17 | Cite each included study and present its characteristics. | P11 Ln 228-235 |
| Risk of bias in studies | 18 | Present assessments of risk of bias for each included study. | P12-13 Ln 264-274 |
| Results of individual studies | 19 | For all outcomes, present, for each study: (a) summary statistics for each group (where appropriate) and (b) an effect estimate and its precision (e.g. confidence/credible interval), ideally using structured tables or plots. | P11-12 Ln 246-261 |
| Results of syntheses | 20a | For each synthesis, briefly summarise the characteristics and risk of bias among contributing studies. | P11 Ln 238-243<br>P12 Ln 264 |
| | 20b | Present results of all statistical syntheses conducted. If meta-analysis was done, present for each the summary estimate and its precision (e.g. confidence/credible interval) and measures of statistical heterogeneity. If comparing groups, describe the direction of the effect. | P13 Ln 278-282, P14 Ln 299-303 |
| | 20c | Present results of all investigations of possible causes of heterogeneity among study results. | P13 Ln 285-294, P14 Ln 305-311 |
| | 20d | Present results of all sensitivity analyses conducted to assess the robustness of the synthesized results. | P13 Ln 293-296, P14 Ln 311-314 |
| Reporting biases | 21 | Present assessments of risk of bias due to missing results (arising from reporting biases) for each synthesis assessed. | P13 Ln 282-284, P14 Ln 303-304 |

| Section and Topic | Item # | Checklist item | Location where item is reported |
|---|---|---|---|
| Certainty of evidence | 22 | Present assessments of certainty (or confidence) in the body of evidence for each outcome assessed. | P15 Ln 327-331 |
| **DISCUSSION** | | | |
| Discussion | 23a | Provide a general interpretation of the results in the context of other evidence. | P15-18 Ln 334-404 |
| | 23b | Discuss any limitations of the evidence included in the review. | P19 Ln 429-433 |
| | 23c | Discuss any limitations of the review processes used. | P19 Ln 421-428 |
| | 23d | Discuss implications of the results for practice, policy, and future research. | P19 Ln 414-420 |
| **OTHER INFORMATION** | | | |
| Registration and protocol | 24a | Provide registration information for the review, including register name and registration number, or state that the review was not registered. | P3 Ln 44-45 |
| | 24b | Indicate where the review protocol can be accessed, or state that a protocol was not prepared. | P5 Ln 107-108 |
| | 24c | Describe and explain any amendments to information provided at registration or in the protocol. | |
| Support | 25 | Describe sources of financial or non-financial support for the review, and the role of the funders or sponsors in the review. | P20 Ln 451-453 |
| Competing interests | 26 | Declare any competing interests of review authors. | |
| Availability of data, code and other materials | 27 | Report which of the following are publicly available and where they can be found: template data collection forms; data extracted from included studies; data used for all analyses; analytic code; any other materials used in the review. | P6 Ln 135-136 |

### Supplemental Method

Search strategy

PubMed

| No. | Search Query | Results |
|---|---|---|
| #1 | "stroke"[MeSH Terms] OR "brain ischemia"[MeSH Terms] OR "cerebral infarction"[MeSH Terms] OR "stroke*"[Title/Abstract] OR "cerebrovascular accident*"[Title/Abstract] OR "brain vascular accident*"[Title/Abstract] OR "Apoplexy"[Title/Abstract] OR "CVA"[Title/Abstract] OR "brain ischemia*"[Title/Abstract] OR "ischemic encephalopath*"[Title/Abstract] OR "cerebral ischemia*"[Title/Abstract] OR "cerebral infarct*"[Title/Abstract] OR "subcortical infarction*"[Title/Abstract] OR "choroidal artery infarction*"[Title/Abstract] | 475,988 |
| #2 | "high intensity interval training"[MeSH Terms] OR "high intensity interval training*"[Title/Abstract] OR "high intensity intermittent exercise*"[Title/Abstract] OR "sprint interval training*"[Title/Abstract] OR "high intensity exercise*"[Title/Abstract] OR "HIIT"[Title/Abstract] OR "HIIE"[Title/Abstract] OR "aerobic interval training*"[Title/Abstract] OR "vigorous aerobic training*"[Title/Abstract] OR "vigorous exercise*"[Title/Abstract] OR "cardiorespiratory fitness training*"[Title/Abstract] | 11,216 |
| #3 | **#1 AND #2** | **331** |
| #4 | "English"[Language] AND 2005/01/01:3000/12/31[Date - Publication] AND (("Randomized Controlled Trial"[Publication Type] OR "randomized"[Title/Abstract] OR "randomised"[Title/Abstract] OR "randomly"[Title/Abstract] OR "trial"[Title/Abstract] OR "placebo"[Title/Abstract]) NOT ("Animals"[MeSH Terms] NOT "Humans"[MeSH Terms])) AND ("Middle Aged"[MeSH Terms] OR "Aged"[MeSH Terms] OR "older adults"[Title/Abstract] OR "elderly"[Title/Abstract] OR "50 years"[Title/Abstract] OR "over 50"[Title/Abstract]) AND (("Humans"[MeSH Terms] OR "Humans"[Title/Abstract] OR "participants"[Title/Abstract] OR "patients"[Title/Abstract]) NOT ("Animals"[MeSH Terms] NOT "Humans"[MeSH Terms])) | **411,428** |
| #5 | **#3 AND #4** | **49** |
| **Total** | | **49** |

Cochrane Library

| No. | Search Query | Results |
|---|---|---|
| #1 | MeSH descriptor: [Stroke] explode all trees | 18,208 |
| #2 | MeSH descriptor: [Brain Ischemia] explode all trees | 5,865 |
| #3 | MeSH descriptor: [Cerebral Infarction] explode all trees | 1,742 |
| #4 | (Stroke* OR 'Cerebrovascular Accident*' OR 'Brain Vascular Accident*' OR Apoplexy OR CVA OR 'Brain Ischemia*' OR 'Ischemic Encephalopath*' OR 'Cerebral Ischemia*' OR 'Cerebral Infarct*' OR 'Subcortical Infarction*' OR 'Choroidal Artery Infarction*'):ti,ab,kw | 87,167 |
| #5 | #1 OR #2 OR #3 OR #4 | 87,415 |
| #6 | MeSH descriptor: [High-Intensity Interval Training] explode all trees | 1,258 |
| #7 | ('High Intensity Interval Training*' OR 'High Intensity Intermittent Exercise*' OR 'Sprint Interval Training*' OR 'High intensity exercise*' OR HIIT OR HIIE OR 'Aerobic interval training*' OR 'Vigorous aerobic training*' OR 'Vigorous exercise*' OR 'Cardiorespiratory fitness training*'):ti,ab,kw | 17,648 |
| #8 | #6 OR #7 | 17,648 |
| #9 | **#5 AND #8** | **803** |
| #10 | English:la | 2,156,075 |
| #11 | ('older adults' OR elderly OR '50 years' OR 'aged 50' OR 'over 50'):ti,ab,kw | 260,444 |
| #12 | MeSH descriptor: [Middle Aged] explode all trees | 415,947 |
| #13 | MeSH descriptor: [Aged] explode all trees | 281,611 |
| #14 | #11 OR #12 OR #13 | 619,556 |
| #15 | MeSH descriptor: [Animals] explode all trees | 897,988 |
| #16 | MeSH descriptor: [Humans] explode all trees | 896,681 |
| #17 | #15 NOT #16 | 1,307 |
| #18 | (humans OR participants OR patients):ti,ab,kw | 1,723,536 |
| #19 | (#16 OR #18) NOT #17 | 1,723,410 |
| #20 | (randomized OR randomised OR 'controlled trial' OR 'clinical trial' OR RCT OR randomly OR placebo):ti,ab,kw | 1,555,168 |
| #21 | **#9 AND #10 AND #14 AND #19 AND #20** | **253** |
| | with Cochrane Library publication date from Jan 2005 to Dec 2025 | 242 |
| **Total** | | **242** |

EMBASE

| No. | Search Query | Results |
|---|---|---|
| #1 | 'cerebrovascular accident'/exp OR 'stroke patient'/exp OR 'brain ischemia'/exp OR 'brain infarction'/exp OR stroke*:ti,ab,kw OR 'cerebrovascular accident*':ti,ab,kw OR 'brain vascular accident*':ti,ab,kw OR apoplexy:ti,ab,kw OR cva:ti,ab,kw OR 'brain ischemia*':ti,ab,kw OR 'ischemic encephalopath*':ti,ab,kw OR 'cerebral ischemia*':ti,ab,kw OR 'cerebral infarct*':ti,ab,kw OR 'subcortical infarction*':ti,ab,kw OR 'choroidal artery infarction*':ti,ab,kw | 889,367 |
| #2 | 'high intensity interval training'/exp OR 'high intensity interval training*':ti,ab,kw OR 'high intensity intermittent exercise*':ti,ab,kw OR 'sprint interval training*':ti,ab,kw OR 'high intensity exercise*':ti,ab,kw OR hiit:ti,ab,kw OR hiie:ti,ab,kw OR 'aerobic interval training*':ti,ab,kw OR 'vigorous aerobic training*':ti,ab,kw OR 'vigorous exercise*':ti,ab,kw OR 'cardiorespiratory fitness training*':ti,ab,kw | 16,197 |
| #3 | **#1 AND #2** | **647** |
| #4 | randomized controlled trial'/de OR 'randomization'/exp OR randomized:ab,ti OR randomised:ab,ti OR rct:ab,ti OR 'controlled trial':ab,ti OR placebo:ab,ti | 1,966,376 |
| #5 | middle aged'/exp OR 'aged'/exp OR 'older adult':ab,ti OR elderly:ab,ti OR '50 years':ab,ti OR 'aged 50':ab,ti | 6,213,368 |
| #6 | **#3 AND #4 AND #5** | **85** |
| #7 | #3 AND #4 AND #5 AND ([middle aged]/lim OR [aged]/lim OR [very elderly]/lim) AND [humans]/lim AND [english]/lim AND [2005-2025]/py | 80 |
| **Total** | | **80** |

Web of Science

| No. | Search Query | Results |
|---|---|---|
| #1 | TS=(Stroke* OR "Cerebrovascular Accident*" OR "Brain Vascular Accident*" OR Apoplexy OR CVA OR "Brain Ischemia*" OR "Ischemic Encephalopath*" OR "Cerebral Ischemia*" OR "Cerebral Infarct*" OR "Subcortical Infarction*" OR "Choroidal Artery Infarction*") | 590,253 |
| #2 | TS=("High Intensity Interval Training*" OR "High Intensity Intermittent Exercise*" OR "Sprint Interval Training*" OR "High intensity exercise*" OR HIIT OR HIIE OR "Aerobic interval training*" OR "Vigorous aerobic training*" OR "Vigorous exercise*" OR "Cardiorespiratory fitness training*") | 14,134 |
| #3 | **#1 AND #2** | **404** |
| #4 | (((LA=(English)) AND PY=(2005-2026)) AND TS=(randomized OR randomised OR randomly OR "controlled trial" OR "clinical trial" OR RCT OR placebo)) AND TS=(humans OR participants OR patients OR elderly OR "older adults" OR "middle aged" OR "50 years" OR "over 50") | 973,604 |
| #5 | **#3 AND #4** | **86** |
| **Total** | | **86** |

<u>Risk of bias assessment</u>

PEDro scores of included studies in this review

| Reference | 1* | 2 | 3 | 4 | 5 | 6 | 7 | 8 | 9 | 10 | 11 | Total (/10) | Quality |
|---|---|---|---|---|---|---|---|---|---|---|---|---|---|
| Boyne, 2016 | Y | 1 | 1 | 0 | 0 | 0 | 1 | 1 | 1 | 1 | 1 | 7 | High |
| Boyne, 2023 | Y | 1 | 1 | 1 | 0 | 0 | 1 | 1 | 1 | 1 | 1 | 8 | High |
| Boyne, 2025 | Y | 1 | 1 | 1 | 0 | 0 | 1 | 1 | 1 | 1 | 1 | 8 | High |
| Do, 2024 | Y | 1 | 0 | 1 | 0 | 0 | 1 | 1 | 1 | 1 | 1 | 7 | High |
| Jin, 2012 | Y | 1 | 0 | 1 | 0 | 0 | 1 | 1 | 0 | 1 | 1 | 6 | High |
| Kim 2025 | Y | 1 | 1 | 1 | 0 | 0 | 1 | 1 | 0 | 1 | 1 | 7 | High |
| Marzolini, 2023 | Y | 1 | 1 | 1 | 0 | 0 | 1 | 1 | 1 | 1 | 1 | 8 | High |
| Moncion, 2024 | Y | 1 | 1 | 1 | 0 | 0 | 1 | 1 | 1 | 1 | 1 | 8 | High |
| Munari, 2018 | Y | 1 | 1 | 1 | 0 | 0 | 1 | 1 | 0 | 1 | 1 | 7 | High |
| Sanberg, 2016 | Y | 1 | 1 | 1 | 0 | 0 | 0 | 1 | 1 | 1 | 1 | 7 | High |
| Soh, 2020 | Y | 1 | 1 | 1 | 0 | 0 | 1 | 0 | 0 | 1 | 1 | 6 | High |

*Not included in the total score; Y, Yes; 1, eligibility criteria; 2, random allocation; 3, concealed allocation; 4, similarity groups at baseline; 5, blinding subjects; 6, blinding therapists; 7, blinding assessors; 8, outcome obtained in >85% of the subjects; 9, intention to treat analysis; 10, between-group statistical comparison; 11, point estimates and measures of variability.

Results of subgroup analysis

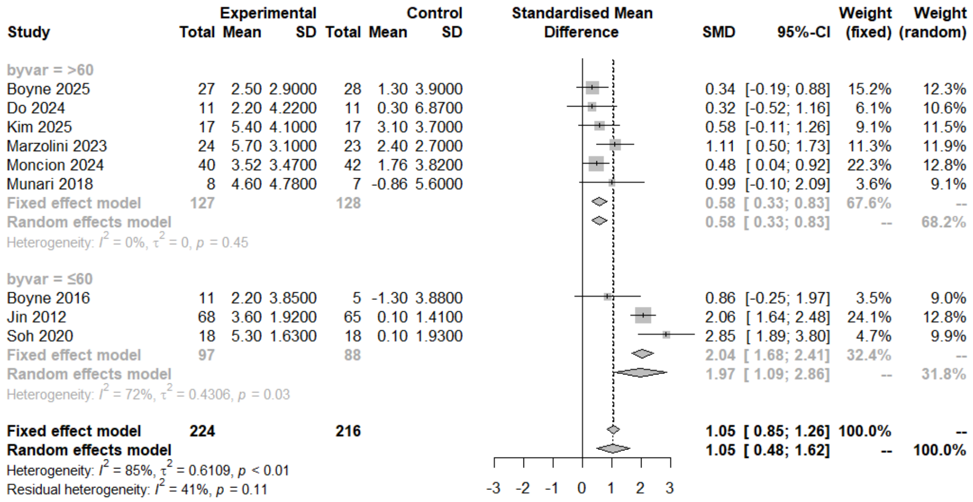


**Figure 1. Subgroup forest plot for VO2peak by Age.** high-intensity interval training demonstrated a more pronounced effect on cardiorespiratory fitness in the ≤60 years group compared to the control group.

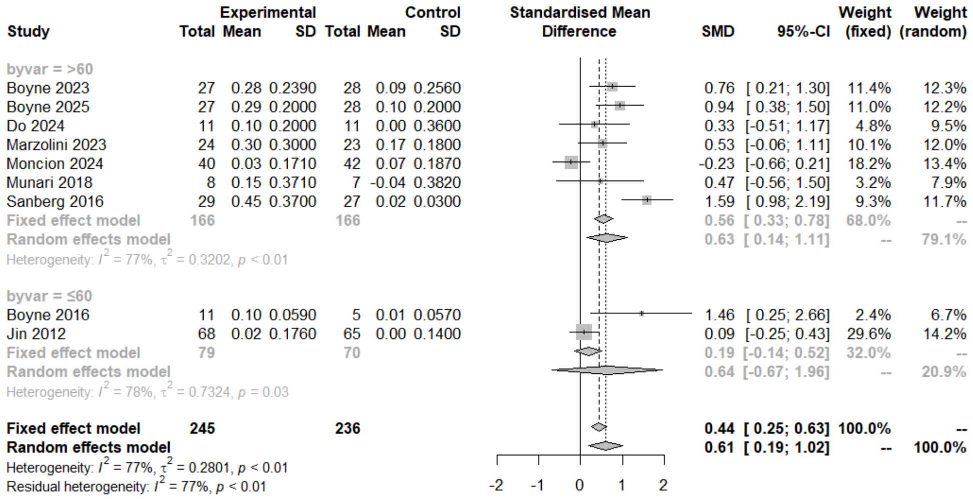


**Figure 2 Subgroup forest plot for gait speed by Age.** The improvement in walking ability following high-intensity interval training did not differ significantly between the age subgroups.

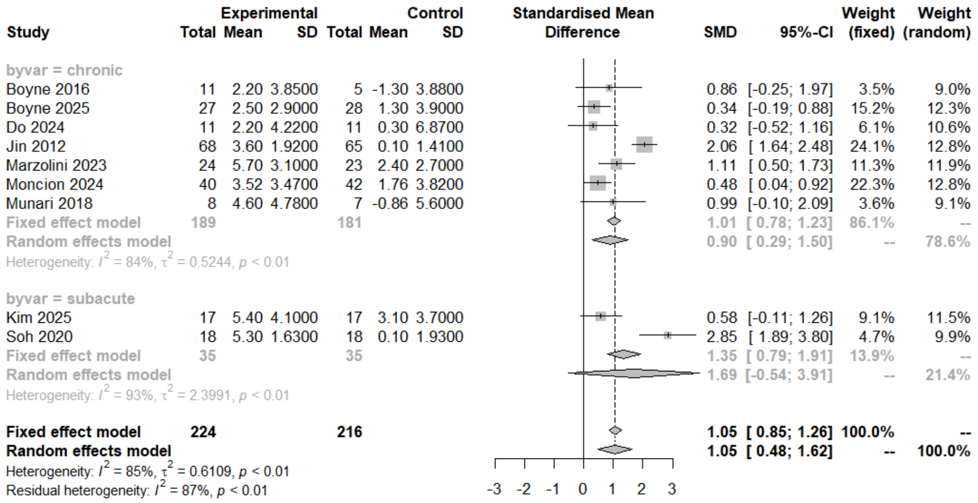


**Figure 3. Subgroup forest plot for VO₂peak by stroke stage.** The improvement in cardiorespiratory fitness following high-intensity interval training did not differ significantly between the onset time of stroke subgroups.

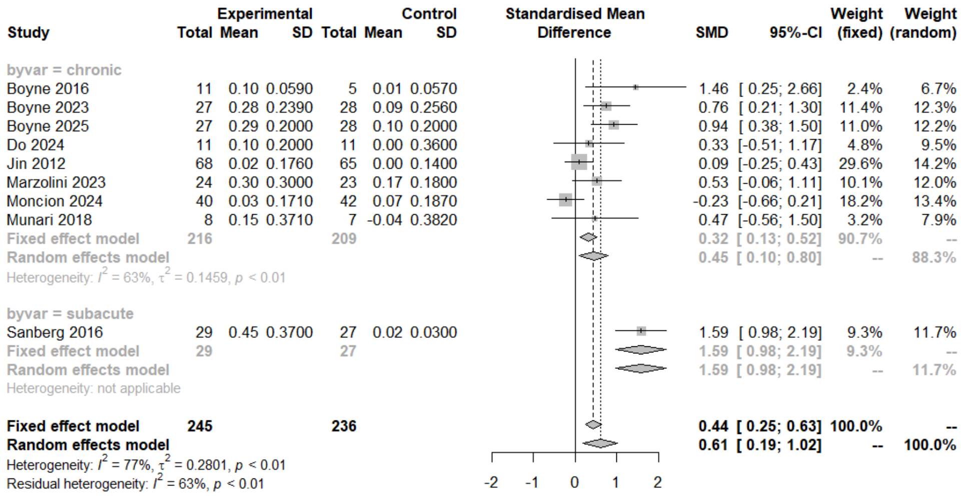


**Figure 4. Subgroup forest plot for gait speed by stroke stage.** high-intensity interval training demonstrated a more pronounced effect on walking ability in the subacute phase compared to the control group.

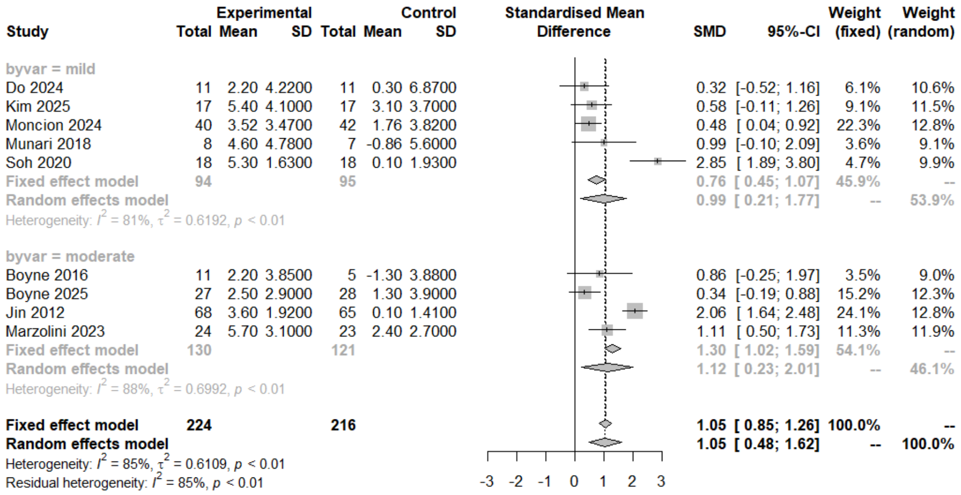


**Figure 5. Subgroup forest plot for VO2peak by stroke severity.** The improvement in cardiorespiratory fitness following high-intensity interval training did not differ significantly between stroke severity subgroups.

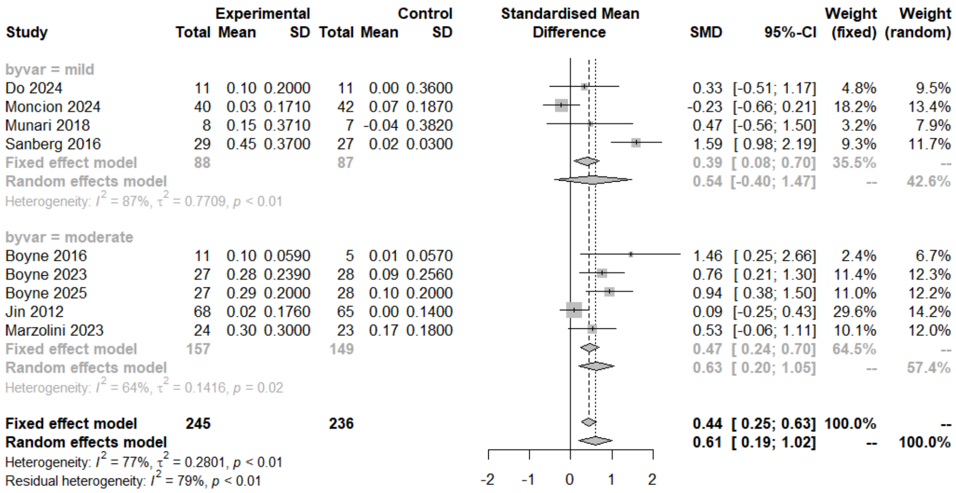


**Figure 6. Subgroup forest plot for gait speed by stroke severity.** The improvement in walking ability following high-intensity interval training did not differ significantly between stroke severity subgroups.

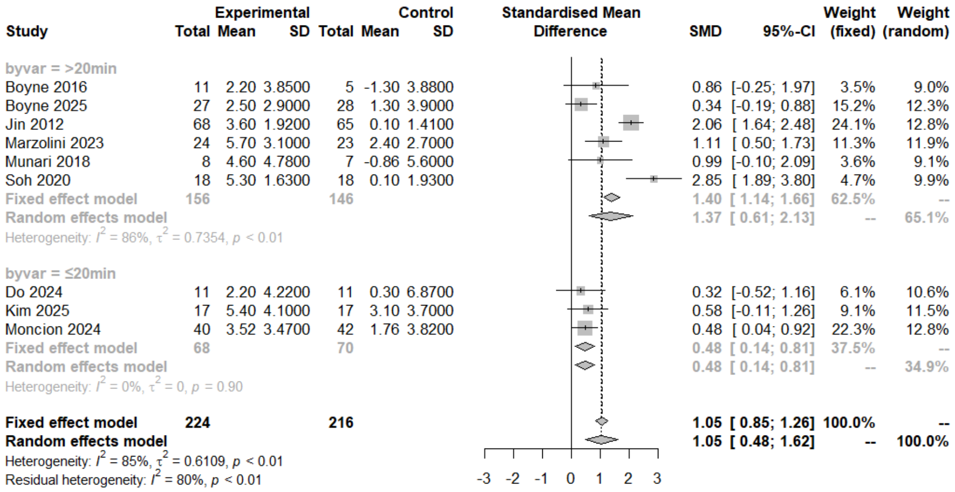


**Figure 7. Subgroup forest plot for VO2peak by intervention time.** The effect of high-intensity interval training on cardiorespiratory fitness was significantly more pronounced in sessions lasting longer than 20 minutes compared to shorter sessions.

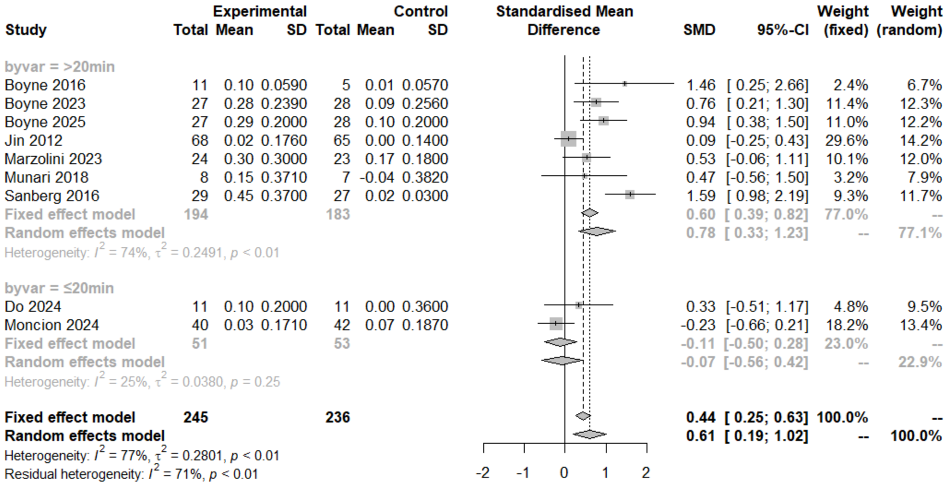


**Figure 8. Subgroup forest plot for gait speed by intervention time.** The effect of high-intensity interval training on walking ability was significantly more pronounced in sessions lasting longer than 20 minutes compared to shorter sessions.

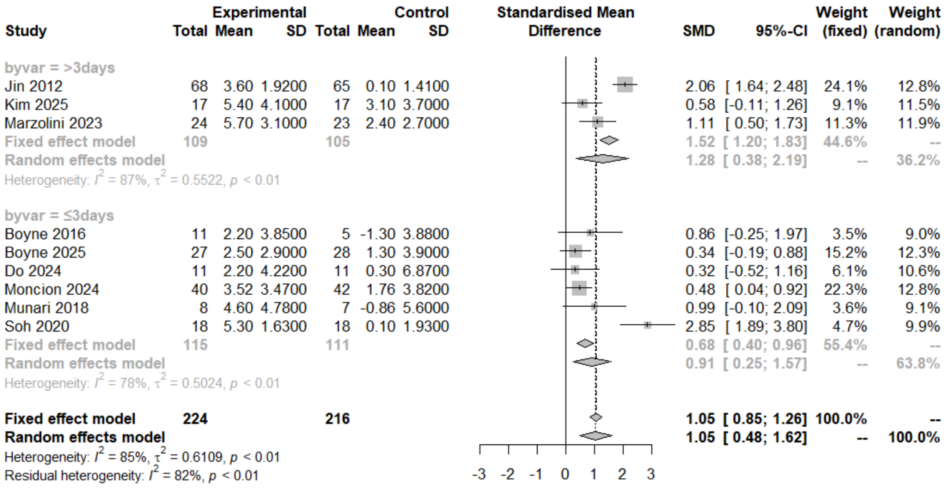


**Figure 9. Subgroup forest plot for VO2peak by intervention frequency.** High-intensity interval training significantly improved cardiorespiratory fitness, and this effect did not differ significantly by intervention frequency.

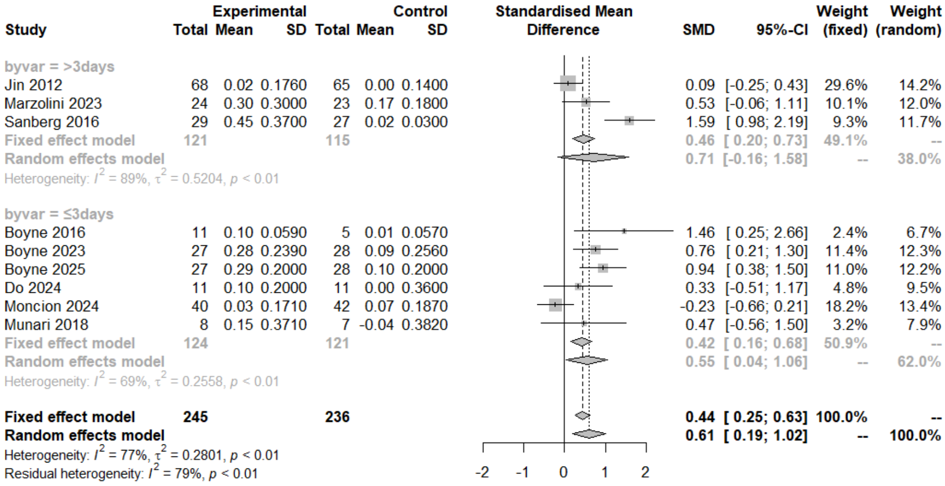


**Figure 10. Subgroup forest plot for gait speed by intervention frequency.** High-intensity interval training significantly improved walking ability, and this effect did not differ significantly by intervention frequency.

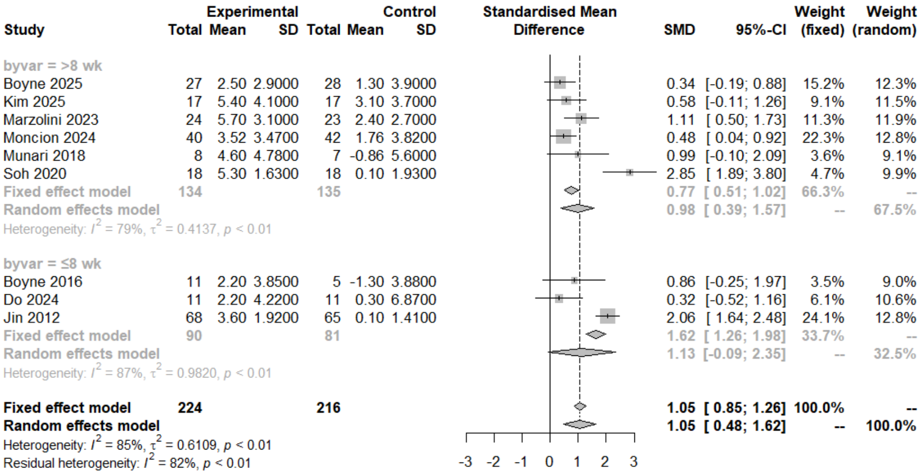


**Figure 11. Subgroup forest plot for $VO_2$peak by intervention length.** High-intensity interval training significantly improved cardiorespiratory fitness, and this effect did not differ significantly by intervention length.

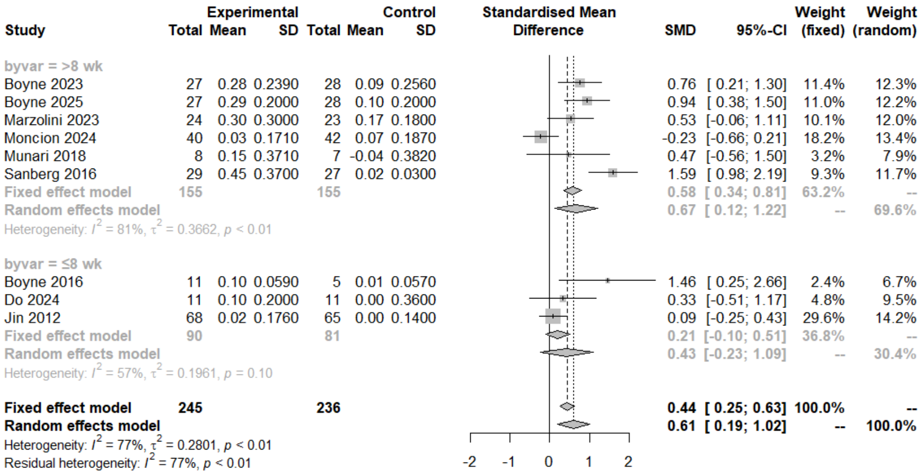


**Figure 12. Subgroup forest plot for gait speed by intervention length.** High-intensity interval training significantly improved walking ability, and this effect did not differ significantly by intervention length.

Sensitivity analysis with Copas selection model

| Outcome | Analysis | SMD | 95% CI | p-value |
|---|---|---|---|---|
| $VO_2$peak | Random effect model | 1.05 | 0.48–1.62 | 0.01 |
| | Copas selection model (adjust) | 1.05 | 0.53–1.57 | 0.01 |
| Gait speed | Random effect model | 0.61 | 0.19–1.02 | 0.01 |
| | Copas selection model (adjust) | 0.60 | 0.21–0.98 | 0.01 |

SMD indicates standardized mean deviation; CI, confidence interval; $VO_2$peak, peak oxygen consumption.

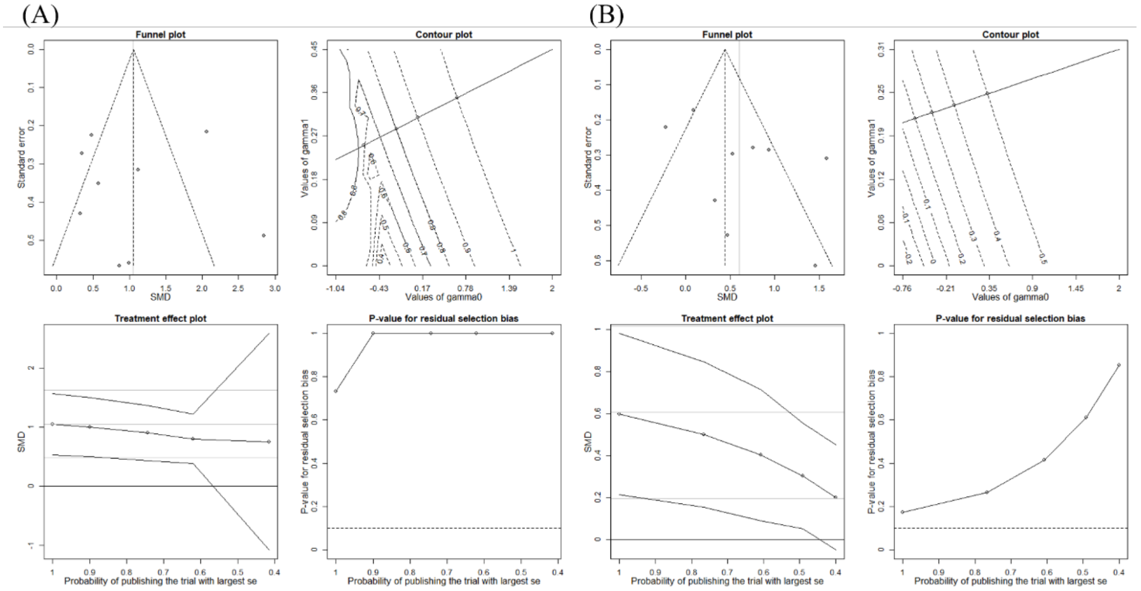


**Figure 13. Results of fitting the Copas model to meta-analysis.** Panel (A) presents the results for cardiorespiratory fitness, and Panel (B) presents the results for walking ability.
